# The distribution of antimicrobial resistance genes across phylogroup, host species and geography in 16,000 publicly-available *E. coli* genomes

**DOI:** 10.1101/2022.08.05.22278465

**Authors:** Elizabeth Pursey, Tatiana Dimitriu, William H. Gaze, Edze R. Westra, Stineke van Houte

**Affiliations:** Environment and Sustainability Institute, Biosciences, University of Exeter, Penryn, Cornwall, United Kingdom; Department of Experimental Medicine, University of Lund, Sweden; European Centre for Environment and Human Health, University of Exeter Medical School, University of Exeter, Penryn, Cornwall, United Kingdom

**Keywords:** antimicrobial resistance, *E. coli*, phylogroups, hosts, regions

## Abstract

*E. coli* is a highly diverse bacterial species that generates a huge global burden of antimicrobial-resistant infections. A wealth of whole genome sequence data is available on public databases for this species, presenting new opportunities to analyse the distribution of antimicrobial resistance (AMR) genes across its genetic and ecological diversity. We extracted and categorised metadata on host species and geographic location and combined this with *in silico* phylogrouping to describe the characteristics of ∼16,000 assembled *E. coli* genomes from the NCBI RefSeq database. We estimated AMR carriage using various metrics: counts of overall genes, multidrug- and extensively drug-resistant categories, and selected β-lactamases of current global concern – *bla*_CTX-M_ and carbapenemase genes. We present estimates of AMR carriage for these metrics by species type (human, agricultural/domestic animal, wild birds and other wild animals), geographic subregion, and across phylogroups. In addition, we describe the distribution of phylogroups within host types and geographic subregions. Our findings show high AMR carriage in commensal-associated phylogroups, agricultural and wild animal hosts and in many subregions. However, we also quantify large biases in sequencing data, the substantial gaps in our knowledge of AMR in many hosts, regions and environmental settings, and the need for systematic sampling to gain a more accurate picture.

## Introduction

*Escherichia coli* is a remarkably diverse species both genetically and ecologically. It occupies various niches, ranging from a commensal of many warm-blooded organisms, to a globally devastating pathogen, to a free-living environmental bacterium [1]. It is a leading cause of mortality associated with drug-resistant infections and was the pathogen responsible for the most deaths attributed to antimicrobial resistance (AMR) in 2019 [2]. In addition, carbapenem-resistant and extended-spectrum β-lactamase (ESBL)-producing *E. coli* are World Health Organization priority pathogens, for which development of new antibiotics is urgently needed [3]. In light of the ongoing AMR crisis, it is necessary to understand how resistance genes are distributed across the ecological and genetic diversity of this species, highlighting the potential risks they pose across contexts.

AMR genes (ARGs) are part of the substantial accessory gene content found in *E. coli* [4], and therefore are not distributed uniformly across the species phylogeny. Acquired ARGs are particularly important due to their extensive ability to transmit horizontally in populations on mobile elements such as plasmids, leading to AMR spread across more distant locations and genetic lineages [5]. Some of these ARGs present significant clinical risks in humans, particularly those with current or potential spread across bacterial host genetic backgrounds, including to other species, and along the commensal-pathogen continuum [6].

Most research has focused on AMR in human-associated and pathogenic isolates [7]. A study focusing on ∼1,000 isolates selected to represent the diversity of *E. coli*, mostly including isolates from non-clinical origins, found ARGs were associated with strains from humans and domesticated animals, independent of phylogroup [8]. Therapeutic and growth promoter usage of antibiotics in agriculture is well-documented, and has been linked to high rates of AMR in livestock-associated isolates [9]. Resistance may also spill over into wild animal populations, and there is increasing evidence of wildlife as potential reservoirs of AMR [10]. A One Health approach to AMR emphasises the interconnectedness of human, animal and environmental microbiomes [11], with the potential for bidirectional flow of ARGs between animal (wild or domesticated), crop, natural and built environment, and human microbiomes. There are also geographic and socioeconomic biases in studies of AMR in *E. coli*. Whilst antibiotics are used most in high-resource settings, the burden of AMR is estimated to disproportionately affect lower and middle income countries, despite the scarcity of data in these regions [2].

A wealth of previous work on *E. coli* has investigated AMR trends in specific settings, such as individual hospitals or farms. As sequencing technologies become more accessible and public genomic databases grow, much larger genomics-based studies of AMR in highly-sampled species like *E. coli* are becoming increasingly feasible. For example, a recent large-scale study of over 70,000 *E. coli* genomes found that the number of distinct ARGs varied between phylogroups, and that specific resistance genes were found more frequently in some, though the pattern of resistance to antibiotic classes remained stable across groups [12]. Another study curated a collection of ∼10,000 *E. coli* isolates sampled from human sources, and identified lineages where >50% of isolates were MDR [13].

In this work we expand on previous large-scale genomic studies of ARGs in *E. coli* by investigating diversity across different host categories and geographic subregions, as well as phylogroups. We characterise a final dataset of ∼16,000 publicly-available assembled *E. coli* genomes, documenting the distribution of ARGs according to several metrics: counts of ARGs, multidrug- and extensively drug-resistant (MDR and XDR) classifications, and presence of clinically-important β-lactamases (*bla*_CTX-M_ and carbapenemases). First, we used model comparison to assess the contributions of host, subregion and phylogroup towards explaining ARG variation in *E. coli*. Then, we used generalised linear models (GLMs) to describe the probabilities of MDR, *bla*_CTX-M_ and carbapenemase presence according to the same predictors. To do so, we used subsampling and resampling approaches to correct for sample size disparities between groupings, and investigate the sensitivity of trends according to subsample sizes.

## Methods

### Genomes

The initial complete dataset of 26,881 *E. coli* genomes was retrieved from the National Center for Biotechnology Information (NCBI) RefSeq database in February 2022 in nucleotide fasta format using ncbi-genome-download v0.3.0 (https://github.com/kblin/ncbi-genome-download).

### Detection of ARGs, phylogrouping and retrieval of metadata

NCBI AMRFinderPlus v3.10.21 was used to predict the identity of acquired ARGs [14]. Clermont phylogrouping [15] was performed *in silico* using the EzClermont command-line tool [16]. Snakemake v6.2.1 [17] was used to generate a pipeline for phylogrouping and detecting ARGs, which was run using the University of Exeter’s Advanced Research Computing facilities. We removed efflux (*acrF*, *emrD* and *mdtM*) and *blaEC* β-lactamase genes from the dataset, as they were either very common or ubiquitous. As we could not use phenotypic resistance categories, we used the classes included in NCBI AMRFinderPlus output as a proxy for categorising genomes as multidrug-resistant (MDR; ARGs conferring resistance to ≥ 3/20 antibiotic classes) and extensively drug-resistant (XDR; ARGs conferring resistance to ≥ 10/20 antibiotic classes) [18].

Host and location metadata were retrieved and categorised using the Bio.Entrez utilities from Biopython v1.77. We filtered genomes for those with complete metadata for host species and geographic location, as well as those not typed as *E. coli* by phylogrouping, which excluded 10,609 genomes, resulting in a final dataset of 16,272 genomes. Geographic locations were split into 20 subregions (Fig. S1) according to Natural Earth data (https://www.naturalearthdata.com/).

All genomes were sorted into the following host species categories: ‘Human’, ‘Agricultural/Domestic animals’, ‘Wild birds’ and ‘Wild animals’. This was achieved using regular expressions constructed by manually reviewing text in the ‘host’ field of the biosample data for each accession number. Wild birds were separated from other wild animals due to their comparatively large numbers of genomes, high mobility and potential for ARG spread [19]. Any animal that is most likely to be associated with humans, such as ‘mouse’ or ‘canine’, was assigned to the ‘Agricultural/Domestic animals’ category, which also included farm animals such as cows and chickens. Broader responses that could not be reliably classified into any specific group, such as ‘Animal’, and ‘Avian’, were not included in further analyses, as well as animals likely to be human food sources such as mussels. Though a conservative approach was taken when assigning genomes to wild animal or bird host groups, these classifications determine the likely category of the host based on species, and cannot rule out cases such as zoo animals.

### Statistical analysis

Data tidying and statistical analysis was done using R v4.0.4, with dplyr v1.0.1, ggplot2 v3.3.6 [20] and MetBrewer [21] for colour palettes. Maps additionally used the R packages sf 1.0-7 [22] and rnaturalearth v0.1.0.

In all cases, binomial GLMs were fitted using glmmTMB v1.1.5 [23]. To assess model fit, simulated residuals were plotted and their dispersion tested using DHARMa v0.4.5 [24]. Prior to all statistical modelling, subgroupings with sample sizes ≤ 100 (e.g. Central Asia) were removed from the dataset. Sample sizes for each predictor in the full dataset are shown in Figure 1A-C.

Initially, maximal models were fitted using the entire dataset with MDR, XDR or *bla*_CTX-M_ genes as a binomial response variable (presence/absence) and all explanatory variables included (host, phylogroup and subregion). For carbapenemase genes, the same response variables were used, but only host and phylogroup were included as predictors due to lack of data coverage for many subregions for these comparatively rare genes. Sample sizes for all combinations of response variable and predictor are shown in Tables S1-3. Next, Akaike information criterion (AIC) scores were used to assess the fit of all possible models using the *dredge* function from MuMIn v1.46.0. In all cases, large ΔAIC values (>50) were seen between the maximal model and the next best model (Table S4), emphasising the contribution of all variables to the distribution of AMR gene measures. Model coefficients for all maximal models are shown in Table S5.

Next, subsampling approaches were used to circumvent disparities in sampling sizes between groups. Separate models were made for each predictor due to the difficulty in 1) taking single representative subsamples spanning all hosts, phylogroups and subregions and 2) estimating means from models with multiple categorical explanatory variables. A subsample was taken for each predictor, giving datasets with equal numbers of genomes from each phylogroup, host or subregion. Several subsample sizes were used to determine the sensitivity of trends to the sample size. Firstly, the dataset was subsampled without replacement down to the lowest group size for each model (e.g. all host categories were sampled to n=234, the sample size for wild birds). In addition, samples with replacement were taken with increasing sizes (n=500, 1000, 2000 and 4000) for hosts, phylogroups and subregions, to investigate the robustness of trends to changing sample sizes. All single predictor models were also run with the full, non-subsampled dataset.

Model estimated means and 95% confidence intervals (CIs) were calculated using *ggpredict* from the ggeffects package [25]. Model coefficients are presented in Table S6 for host, Table S7 for subregion and Table S8 for phylogroup. We did not perform post-hoc comparisons or report *P*-values in the manuscript due to the large number of pairwise comparisons that would be necessary (e.g. 91 comparisons for each model of subregion data). In addition, we did not have *a priori* hypotheses about group differences to test, but rather aimed to describe the patterns in the data with our modelling approaches. Finally, many groups had large sample sizes, which can produce a significant result even for a biologically insignificant effect size [26, 27].

## Results

### Quantification of sample sizes across hosts, phylogroups and subregions in the RefSeq dataset

Initially, we characterised the RefSeq dataset by looking at sample sizes across phylogroups, host categories, and geographic subregions (Fig. 1A-C, Tables S1-3). Phylogroups B1, A and B2 collectively made up 74% (12,109/16,272) of genomes, with smaller numbers found across C, E, F, G and U (Fig. 1A). Sixty-five genomes were typed as U/cryptic or cryptic. The majority of genomes had a sampling location in North America (n=5,202) or Eastern Asia (n=3,790), whilst many regions were represented poorly such as Micronesia, Melanesia and Polynesia (n=1 for all), Central Asia and Caribbean (n=3 for both) and Middle Africa (n=10) (Fig. 1B). Finally, 96% of the *E. coli* genomes were isolated from humans and agricultural or domestic animals (15,549/16,272) with only 234 from wild birds and 489 from other wild animals (Fig. 1C).

**Figure 1.**
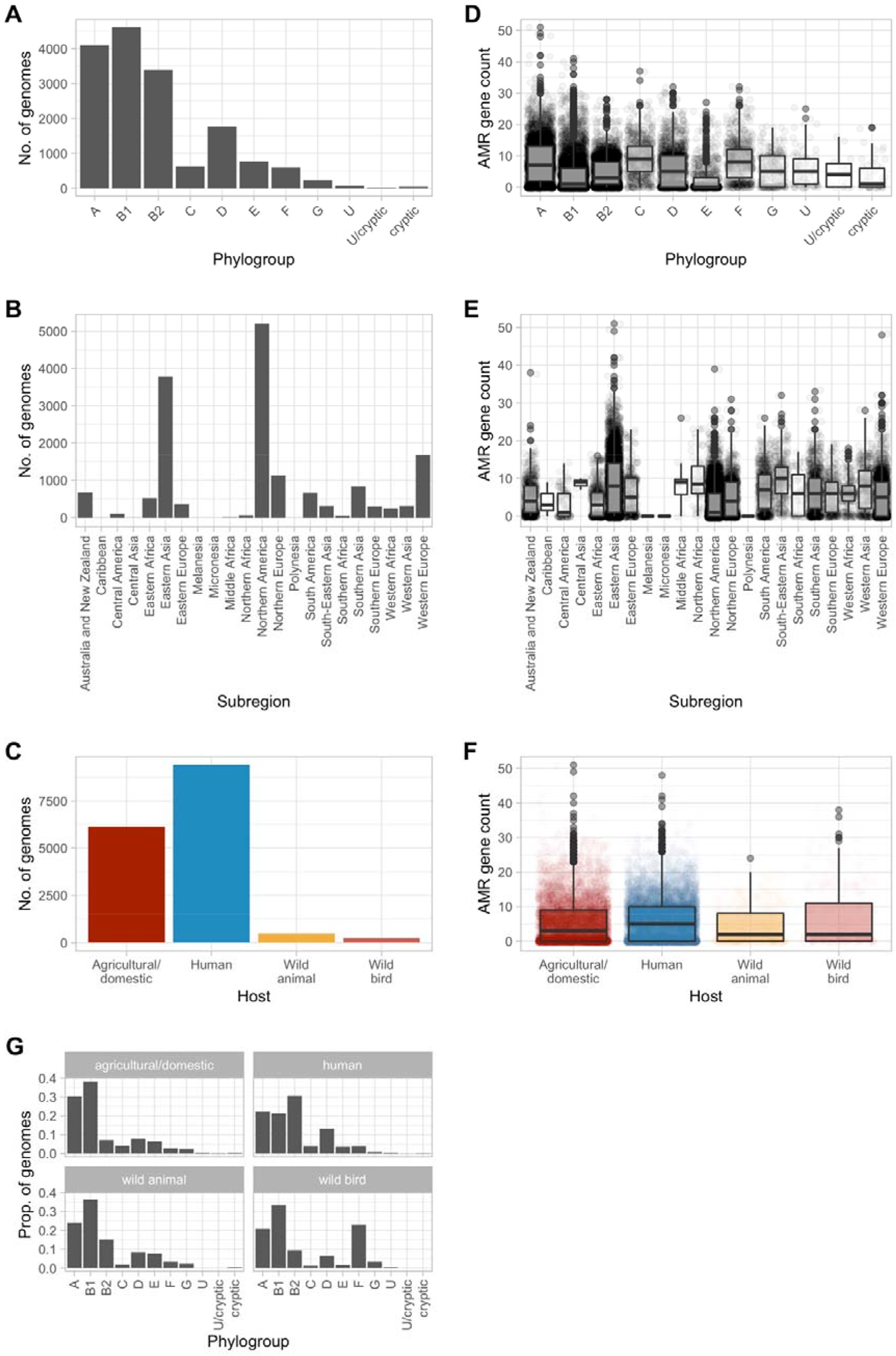
Total number of genomes per A) phylogroup, B) geographic subregion and C) host species category. Counts of ARGs for D) phylogroup, E) geographic subregion and F) host species category, with raw data plotted below boxplots. G) Proportion of genomes in each phylogroup per host category.

### ARG counts across the diversity of *E. coli* sequences

We measured counts of ARGs detected across the diversity of genomes to get an overall measure of how AMR carriage was distributed across the dataset (Fig. 1D-F). Overall, the mean number of ARGs per genome for the entire dataset was 5.75. Median ARG counts were highest in phylogroups C, F and A (9, 8 and 7 respectively; Fig. 1D). Meanwhile the lowest median counts were in groups E (0) and B1 (1). A wide range of ARG counts was also seen between and within subregions (Fig. 1E). Among the most well-sampled regions, genomes from Eastern Asia had the highest median ARG count (8), though relatively high numbers were also seen in Western Europe (median ARG count = 5). Additionally, higher median ARG numbers were seen in less represented regions such as Northern (8.5), Middle (9) and Southern Africa (6) and Central Asia (9). High within-group diversity in ARG counts was again seen for host categories, with highest median counts in genomes from humans (5) and agricultural animals (3).

### Hosts and subregions show different phylogroup patterns

We investigated the distribution of *E. coli* phylogroups within hosts (Fig. 1G) and geographic subregions (Fig. S2). *E. coli* isolates from agricultural and domestic animals mostly belonged to phylogroups A and B1 (n=1,863 and n=2,354, respectively). Those from humans were most likely to belong to phylogroup B2 (n=2,870), as well as A and B1 (n=2,071 and n=2,000 respectively). There were also comparatively high numbers of phylogroup D isolates in genomes from human hosts (n=1,237). A higher proportion of phylogroup F isolates were seen in wild birds (n=54), whilst wild animals had slightly higher proportions of B2 isolates (n=74) than agricultural or domesticated animals.

The frequencies of different phylogroups varied between geographic subregions (Fig. S2). While A, B1 and B2 were typically the largest groups, their relative proportions varied. For example, B1 was more common in Eastern (n=195) and Western (n=94) Africa, B2 was highest in Australia and New Zealand (n=216) and Eastern Europe (n=112), while A was most common in South-Eastern (n=131) and Western (n=117) Asia. Isolates belonging to phylogroup D were also relatively common, with notably higher proportions in regions including Northern Africa (n=17) and Eastern Europe (n=86).

### Specific measures of AMR carriage

Following characterisation of the dataset, we began analysing more specific measures of AMR carriage. First, we categorised genomes as MDR (≥ 1 ARGs conferring resistance to ≥ 3 classes) and XDR (≥ 1 ARGs conferring resistance to ≥ 10 classes) following detection of 402 unique resistance genes conferring predicted resistance to 19 classes. We also looked at the presence of selected β-lactamase genes of current global health concern. We detected 40 unique *bla*_CTX-M_ genes across the final dataset, most commonly *bla*_CTX-M-15_ (n=2,536), *bla*_CTX-M-14_ (n=1,063) and *bla*_CTX-M-55_ (n=903). Meanwhile, a total of 46 different carbapenemases from the NDM, KPC, OXA, IMP, VIM, IMI and GES classes were identified, the most frequent being NDM-5 (n=893), KPC-2 (n=264), NDM-1 (n=219) and OXA-48 (n=189).

We generated model estimated mean proportions with 95% CIs for models of MDR (Fig. 2), XDR (Fig. S3) and *bla*_CTX-M_ (Fig. 3) presence, according to host, phylogroup and subregion for both the full dataset, and a dataset subsampled down to the smallest group size without replacement to adjust for large sample size differences between groups. The same process was repeated for carbapenemase gene presence (Fig. 4), but subregion was not used as a predictor due the rarity of carbapenemase-positive genomes across many regions. Trends were broadly consistent across the two datasets and estimates for the full dataset are quoted in the subsequent text.

#### 1. MDR and XDR

MDR genomes were common, with a given genome from the majority of phylogroups, subregions and hosts having > 0.4 probability of being MDR (Fig. 2). Within phylogroups, multidrug resistance was most common in groups C (0.83, 95% CI=0.80-0.86), F (0.75, 95% CI=0.71-0.78) and A (0.72, 95% CI=0.70-0.73), whilst being comparatively uncommon in group E isolates (0.25, 95% CI=0.22-0.28). Isolates from humans (0.60, 95% CI=0.58-0.60) and agricultural/domestic hosts (0.51, 95% CI=0.50-0.52) had the highest probabilities of MDR, though estimates from wild animals (0.44, 95% CI=0.39-0.48) and birds (0.43, 95% CI= 0.37-0.50) were reasonably high. Finally, South-Eastern Asia (0.86, 95% CI = 0.83-0.90) and Western Africa (0.78, 95% CI=0.15-0.72) had the highest estimated proportions of MDR genomes, while the lowest were in Northern America (0.40, 95% CI=0.38-0.41) and Central America (0.42, 95% CI=0.33-0.52).

XDR genomes were less common, and not detected at high enough frequencies to estimate their probabilities in the minimum subsampled dataset for phylogroup B2 and wild animal hosts (Fig. S3). The phylogroups with the highest probabilities of XDR were A (0.092, 95% CI=0.083-0.10) and F (0.07, 95% CI=0.052-0.093), whilst remaining very low for B2 (0.003, 95% CI=0.0018-0.0058) and E (0.018, 95% CI=0.012-0.03). The highest XDR probabilities were estimated in wild birds (0.13, 95% CI=0.091-0.18), with probabilities below 0.06 for other host categories. Finally, XDR genomes were by far the most likely to occur in Eastern Asia (0.16, 95% CI=0.14-0.17), and below 0.08 for all other regions.

**Figure 2.**
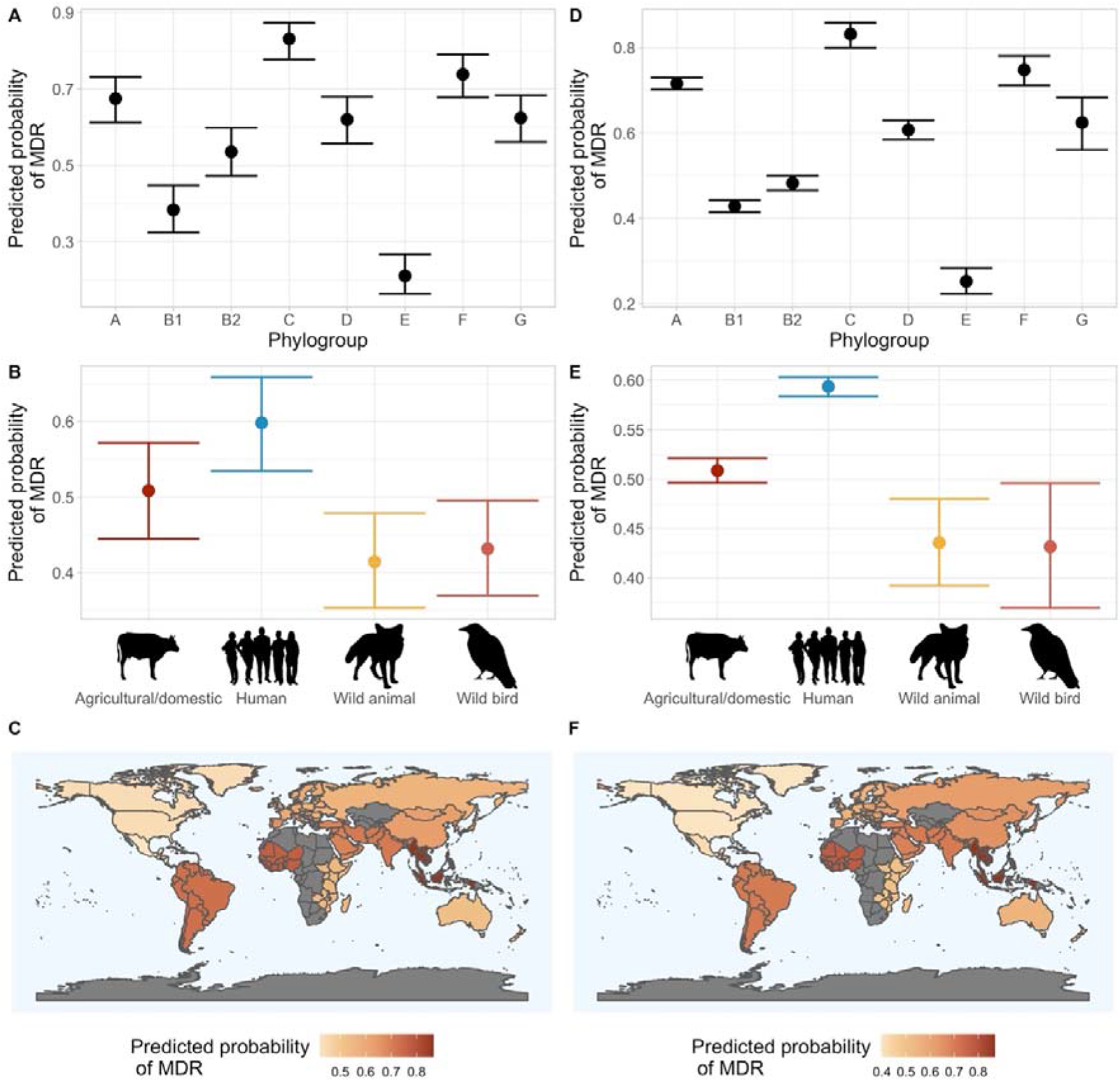
Estimated mean probabilities and 95% CIs of MDR from binomial GLMs for datasets subsampled without replacement to the minimum group size for A) phylogroup, B) host category and geographic subregion. The same model types for the full non-subsampled dataset for D) phylogroup, E) host category and F) geographic subregion. CIs for subregion models can be found in Figure S6.

#### 2. bla_CTX-M_ and carbapenemase genes

The presence of the *bla*_CTX-M_ class of β-lactamases was highly variable between groups (Fig. 3). Estimated probabilities ranged from 0.08 (95% CI = 0.066-0.11) in phylogroup E to 0.51 (95% CI=0.47-0.55) in phylogroup F, whilst *bla*_CTX-M_ genes were more associated with human hosts (0.33, 95% CI=0.32-0.34) than other host categories. The probabilities of *bla*_CTX-M_ genes being present also varied widely between subregions, with the highest in South-Eastern Asia (0.59, 95% CI=0.53-0.64) and Eastern Europe (0.52, 95% CI=0.47-0.57), and all other subregions below 0.45.

**Figure 3.**
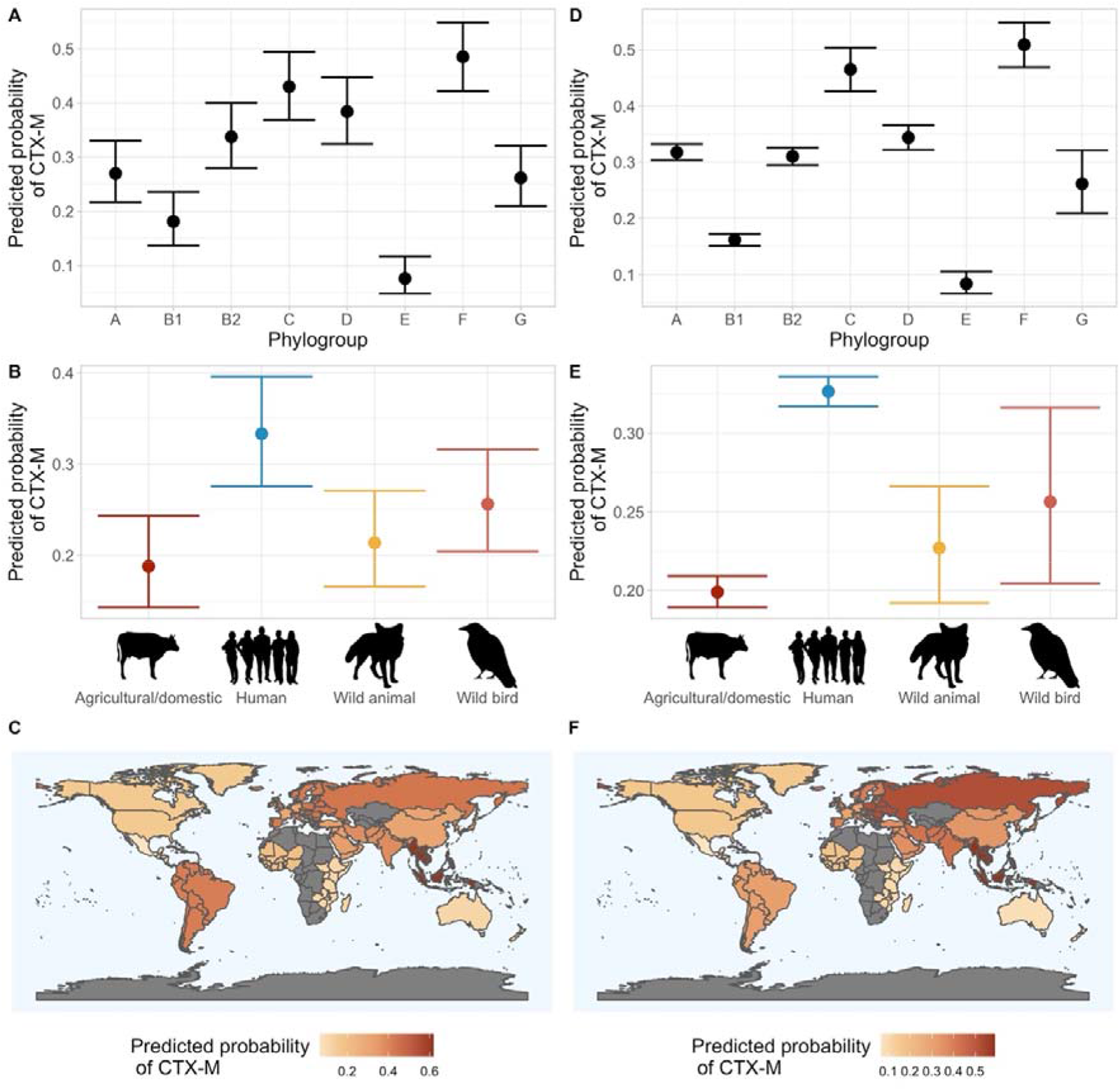
Estimated mean probabilities and 95% CIs of *bla*_CTX-M_ presence from binomial GLMs for datasets subsampled without replacement to the minimum group size for A) phylogroup, B) host category and C) geographic subregion. The same model types for the full non-subsampled dataset for phylogroup, E) host category and F) geographic subregion. CIs for subregion models can be found in Figure S12.

Carbapenamase genes were far less common than *bla*_CTX-M_ genes (Fig. 4). There was a clear split between phylogroups C, F, A and D, which were more likely to possess carbapenemases (probabilities >0.15), and the remaining groups which had probabilities of 0.05 or below. The highest proportions of carbapenemases were estimated in wild bird (0.25, 95% CI=0.20-0.31) and human (0.14, 95% CI= 0.13-0.15) hosts.

We further investigated sampling bias for carbapenemase-positive isolates in wild birds due to the high proportion of genomes possessing these genes. Isolates from wild birds possessing carbapenemases were only represented in 3 subregions: Australia and New Zealand (n=25), Eastern Asia (n=32) and Western Asia (n=1). Sequencing bias was evident; for example, 68 out of 234 wild bird isolates were from one study of silver gulls in Australia (which made up 68 out of the 93 isolates from this region). This study (PRJNA630096) specifically sought and sequenced isolates conferring resistance to critically important antimicrobials. Further carbapenemase-producing isolates came from studies in China (PRJNA669620 and PRJNA349231) that specifically studied carbapenem-resistant *E. coli*.

**Figure 4.**
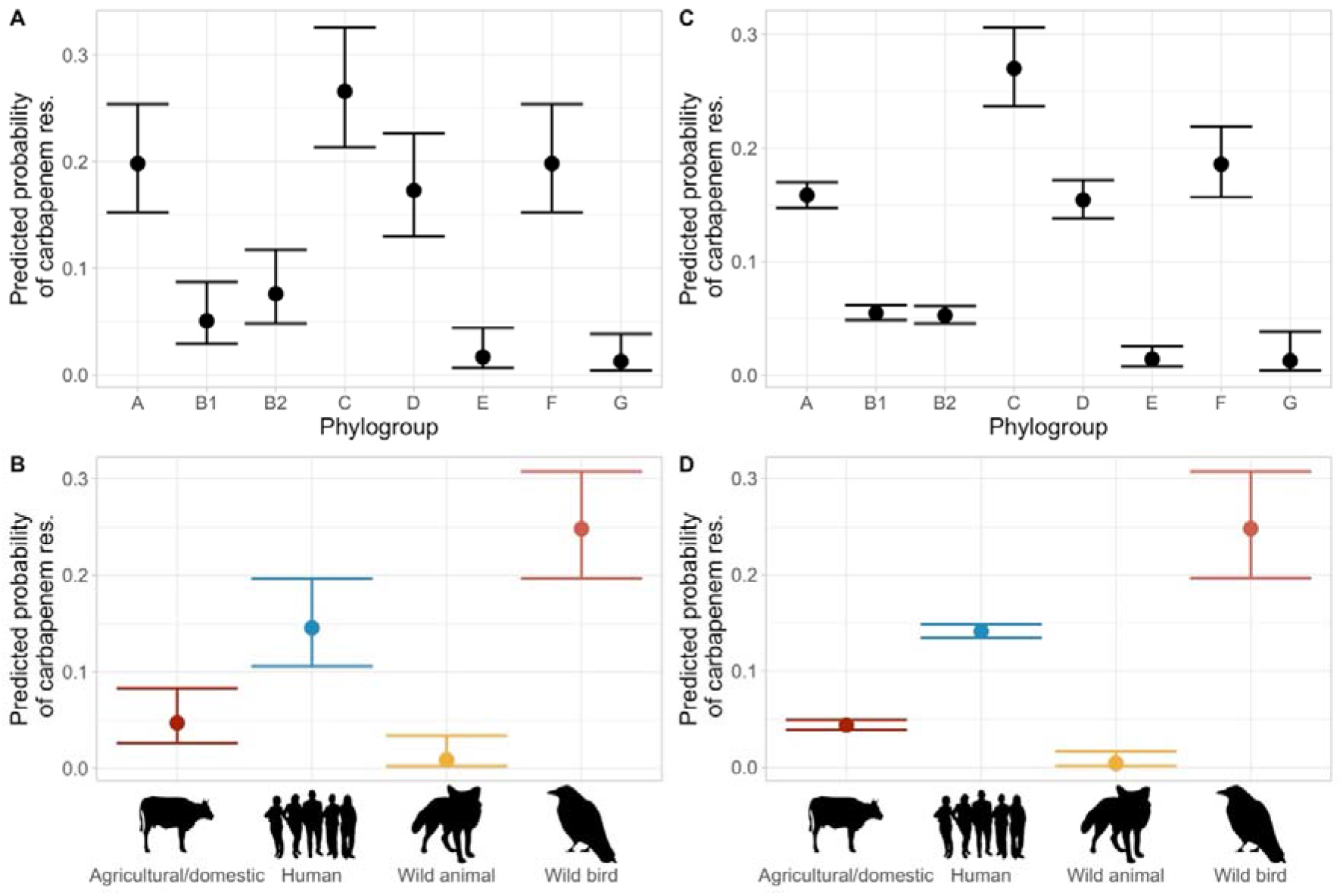
Estimated mean probabilities and 95% CIs of carbapenemase presence from binomial GLMs for datasets subsampled without replacement to the minimum group size for A) phylogroup and B) host category. The same model types for the full non-subsampled dataset for C) phylogroup and D) host category.

### Subsampling and resampling approaches do not alter major trends

Due to the large discrepancies in sample size between hosts, regions and phylogroups in this dataset, we used additional sampling with replacement to examine the sensitivity of the observed trends to changing sample sizes of 500, 1000, 2000 and 4000 (Fig. S4-S12). This showed that, although fluctuations in estimated proportions occurred (as well as expected reductions in CIs with increased sample size), broader trends were reproducible across wide variations in subsample size. In some smaller subsamples, insufficient genomes possessed the metric of AMR carriage (e.g. a carbapenemase gene) to make estimates in some subgroupings.

## Discussion

Though we are in the era of large-scale genomics studies, with thousands of genomes available for species such as *E. coli*, it remains challenging to collate this information to answer questions about AMR prevalence. In this study, we characterised the RefSeq *E. coli* dataset for which metadata was available, investigating sampling bias and AMR carriage according to various metrics for different hosts, phylogroups and geographic subregions.

### How evenly sampled are publicly-available E. coli genomes?

In first characterising this dataset, large sample size discrepancies in the locations and hosts *E. coli* genomes were sourced from were apparent. This was not unexpected. Previous large-scale work on *E. coli* genomes isolated from human hosts has emphasised the sampling biases towards clinically-important isolates and lineages, as well as high-income countries, even in the most well-sampled host species [13]. Other work has used literature searches to quantify how poorly sampled wild animals are for *E. coli*, with small within-group sample sizes in the few studies available [28]. Our data adds to this body of knowledge emphasising how little is known about ARG burden across the diversity of this species, despite the vast amount of sequencing data available.

We investigated the effects of subsampling all groups to the minimum sample size, as well as resampling up to larger sample sizes. This approach cannot replace unbiased sampling, particularly for those groups that are less well represented. However, it is interesting to note that estimates of AMR carriage for larger groups, such as phylogroup A or human hosts, did not alter substantially regardless of the subsampling regime. Additionally, the spread of counts of ARGs was wide in these groups, indicating that they were not necessarily dominated by one or few lineages.

Previous work has also investigated the potential effect of sampling bias on AMR estimates using *E. coli* genomes from GenBank [29]. These authors found that 1) the average distance of a given newly sequenced *E. coli* genome is equivalent to that between known close relatives, such as O157:H7 genomes, and 2) the results of a population genetics analysis of AMR did not change substantially when all human or unknown source genomes were removed, or all genomes for which the GenBank record referenced antibiotic resistance. Therefore, it is possible that, for more well-sampled subgroupings of *E. coli*, our current sequence collection already spans a high diversity of this species.

### Phylogroup distributions within hosts and subregions

Though the most common phylogroups were A, B1 and B2, their proportions varied substantially between hosts. Phylogroup B1 was the most frequent in agricultural and wild animals. This is broadly consistent with many smaller-scale studies for which B1 was the most frequent in agricultural settings. For example, B1 made up 71% of agriculture-associated isolates in a study in the Philippines [30], 50% of those in a study of sheep farming in China [31], and 35% of isolates from cattle and their attendants in Tanzania [32]. The wide variation in actual proportions observed in these different studies may be due to smaller sample sizes (n=17-100 in those referenced), as well as location and species effects. Human isolates were proportionally more likely to be B2 than those from other hosts. *E. coli* isolates from group B2 tend to possess more virulence traits than those from A and B1, though this virulence is proposed to be a by-product of commensalism [33]. Geographic differences in predominant commensal *E. coli* phylogroups have broadly represented a shift from A to B2 in industrialised nations over the past decades, with these being the most common phylogroups in humans [34]. We saw this trend to some extent – European regions and Australia and New Zealand had high proportions of B2. Contrastingly, we found that other regions such as North America and Eastern Asia had B1 as the predominant phylogroup.

### Estimating AMR carriage across phylogroups, hosts and subregions

We used various metrics of AMR carriage to get an overview of ARG distribution in publicly-available *E. coli* genomes. When comparing phylogroups, A, C and F were repeatedly associated with more ARGs as well as clinically-important β-lactamases. Phylogroup A is a major group that appears to be more generalist. It is spread across vertebrate hosts [8], and contains mostly human commensal strains [35] as well as laboratory strains [36]. Although the presence of engineered strains may partly explain the high AMR metrics in this group, our results also imply that ARGs may be frequent in commensal lineages. Phylogroups commonly present in the human gut could be a reservoir for resistance genes and plasmids [37], potentially allowing their horizontal spread into other, more problematic lineages or species.

As they are minor phylogroups, comparatively less is known about groups C and F. In terms of pathogenic potential, the sequence type complex 88 (STc88) lineage of group C is one of the main avian pathogenic *E. coli* (APEC) strains [38]. This phylogroup also includes ST410, a recently-emerged MDR lineage with fluoroquinolone resistance, ESBL (*bla*_CTX-M-15_) and carbapenemase (*bla*_OXA-181_ and *bla*_NDM-5_) genes [38]. Meanwhile, phylogroup F has been associated with fewer virulence traits than closely related groups [39], though it has also been linked to extraintestinal pathogenic (ExPEC) infections [38] and contains the MDR STc648 lineage [40]. Therefore, these phylogroups could represent highly virulent and/or drug-resistant lineages and may warrant further investigation.

Unsurprisingly, human hosts were associated with more ARGs than other host categories. However, high levels were also seen in agricultural and domestic animal species for some measures, such as MDR. In addition, *bla*_CTX-M_ and carbapenemase genes were detected in all the animal host categories to some degree. Antibiotics are still extensively used in animal agriculture, selecting for ARGs both directly in the gut and through excretion of antibiotics into soil and the local environment [41]. Despite this there is no standardised global surveillance system used in animal agriculture that is equivalent to those used in humans [41]. There is evidence for transmission of *bla*_CTX-M_ variants in *E. coli* from humans to farmed animals and the environment, which also occurs in the reverse direction via food products [42]. Only limited studies document the occurrence of carbapenemases in *E. coli* strains from wildlife, agricultural animals and soils, though they have been reported in these contexts [43, 44]. This highlights the need for further prevalence studies to determine the extent to which these genes are spreading in different environments.

The proportionally high numbers of carbapenemase-possessing isolates (and potentially XDR isolates) from wild birds in this work are likely due to studies that enriched for ARGs. However, this emphasizes our lack of knowledge on the true extent of AMR spread in the natural world. High levels of problematic ARGs in wild birds, particularly generalist species, could occur due to their relatively high mobility compared to other taxa, and their utilisation of diverse foraging habitats, both of which increase potential exposure to anthropogenic sources of resistant bacteria [45, 46].

Finally, estimates of ARG prevalence varied geographically depending on the metric used. The highest AMR carriage was generally estimated for Eastern and South-Eastern Asia across measures. However, other regions were particularly high for individual measures. For example, Eastern Europe had high levels of *bla*_CTX-M_ genes and MDR genomes were more common in Western Africa. These trends could either reflect genuine differences in AMR levels and individual gene prevalence, or regional differences in sampling strategies and genomic surveillance. There are large gaps in AMR surveillance for low and middle income (LMIC) countries. A recent study utilised the relationship between socioeconomic characteristics and AMR prevalence to model AMR levels for underrepresented countries, estimating high levels of third-generation cephalosporin resistance in *E. coli* from Western Asia [47]. Genomic surveillance has been leveraged during the COVID-19 pandemic and for other infectious diseases, but next-generation sequencing capacity is low in many regions [48]. However, the development of more many sequence analysis tools to detect ARGs [49], as well as developing capacity to incorporate whole genome sequencing into AMR surveillance in LMICs [50], may lead to this becoming a powerful tool in the future.

### Public genomic databases – limitations, benefits and future work

We show the void in sequencing data from *E. coli* isolates outside of human and agricultural settings in a very large dataset, as well as stark regional disparities, with scarce publicly-available data representing the majority of global regions. Whilst this study characterises this currently available dataset, representative estimates of ARG burden in *E. coli* sequences cannot be made without systematic, representative sampling. Notably, it is concerning that ARGs that inactivate our last-resort antibiotics are being detected and potentially spread in any environmental context, and that we do not have sufficient genomic data to investigate this. Despite their limitations, publicly-available datasets are important, allowing access to sequencing data for scientists and the public across the globe, as well as improving reproducibility. Future systematic sampling and genomic surveillance in previously under-investigated settings will give a more accurate picture of the scale of the AMR problem.

## Data Availability

All code used to generate and analyse data are available at https://github.com/elliekpursey/AMR-Ecoli

## Author contributions

EP, SVH and ERW conceptualised the study. WHG refined the directions of the analysis and methodology. EP performed all data and statistical analysis and wrote the first manuscript draft. All authors contributed to interpretation of the results and editing subsequent versions of the manuscript.

## Acknowledgements and funding

EP was supported by a PhD studentship equally funded by a grant from the European Research Council under the European Union’s Horizon 2020 research and innovation programme (ERC-STG-2016-714478 to ERW) and the College of Life and Environmental Sciences, University of Exeter, UK. EP thanks Liam Langley for statistical modelling advice, and Elze Hesse and Kate Baker for feedback on the analyses. TD is supported by European Research Council (ERC-2017-ADG-788405 to ERW). SVH acknowledges BBSRC for funding (BB/R010781/1 and BB/S017674/1).

