## Supplementary material for "The distribution of antimicrobial resistance genes across phylogroup, host species and geography in 16,000 publicly-available *E. coli* genomes"

**Supplementary Figures**

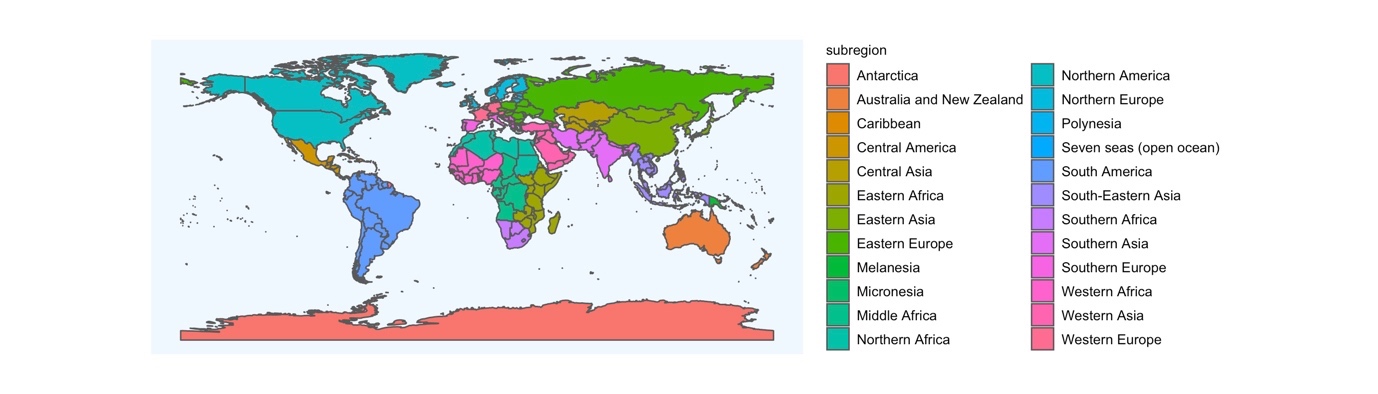

Figure S1. Map showing subregion classifications used in the study.

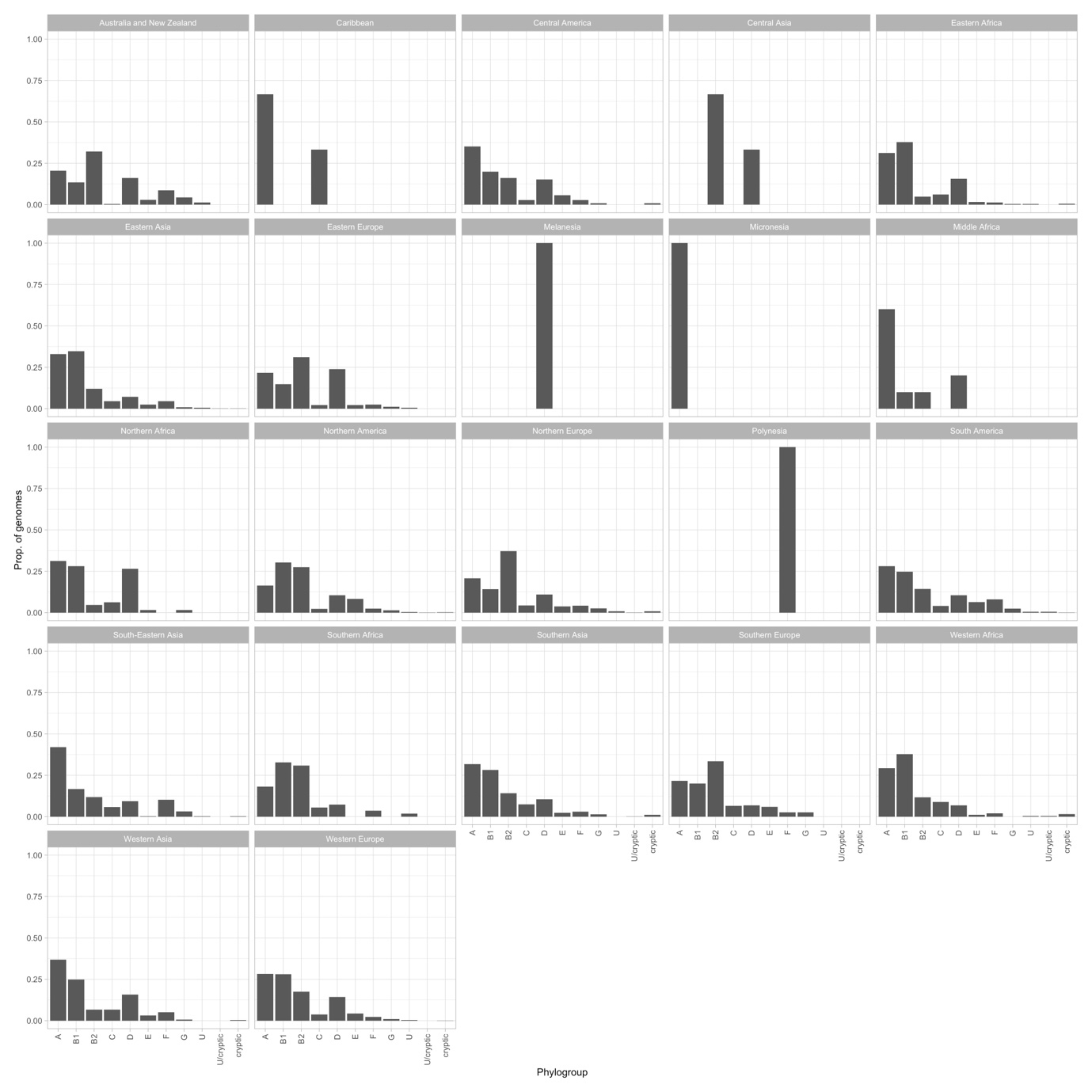

Figure S2. Proportion of genomes in each phylogroup per geographic subregion

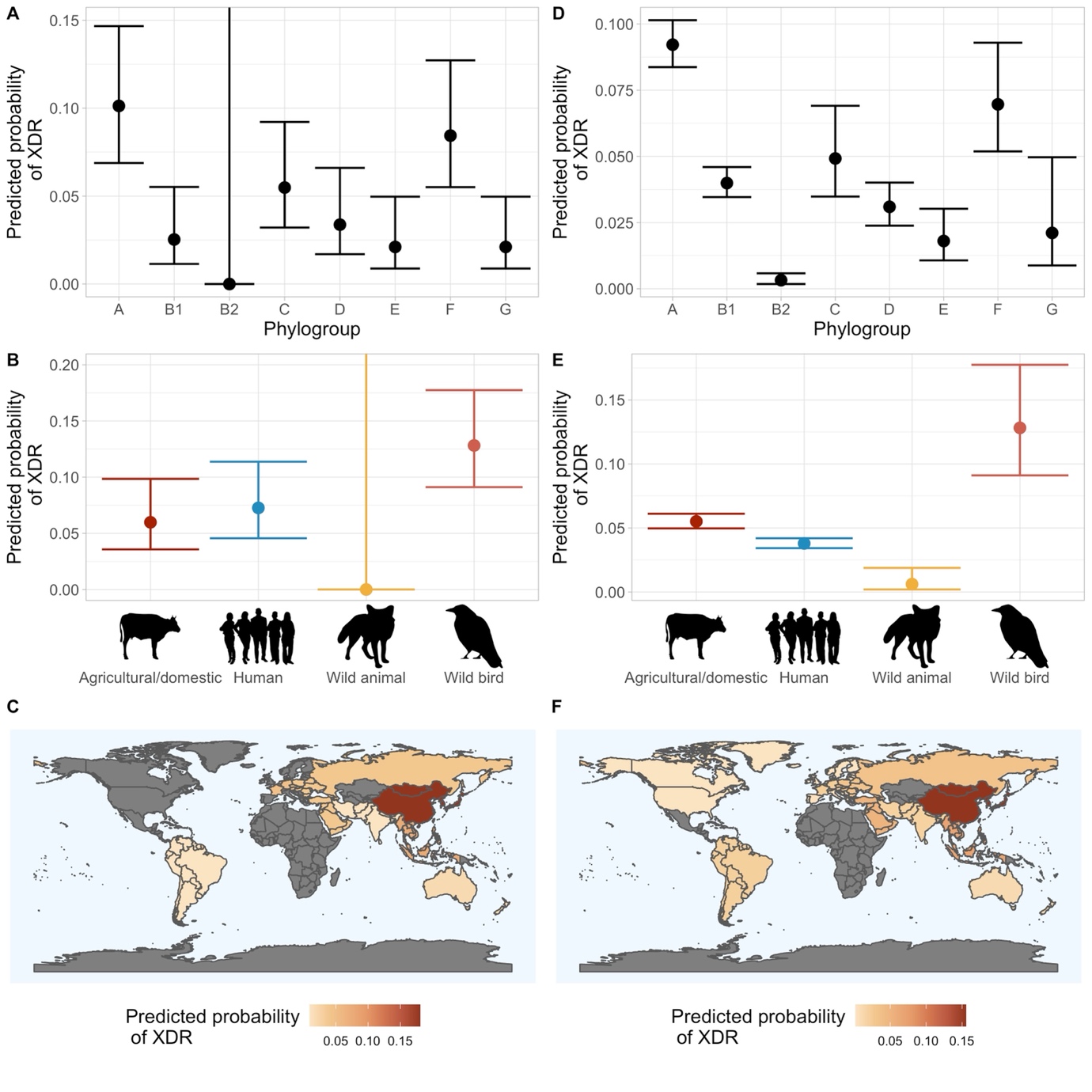

Figure S3. Estimated mean probabilities and 95% CIs of XDR from binomial GLMs for datasets subsampled without replacement to the minimum group size for A) phylogroup, B) host category and C) geographic subregion. The same model types for the full non-subsampled dataset for D) phylogroup, E) host category and F) geographic subregion. CIs for subregion models can be found in Figure S9. Where CIs span the full range, there was insufficient data to estimate the probability for this level of the predictor. Subregions where CIs spanned the full range have been removed from map plots (shown in grey).

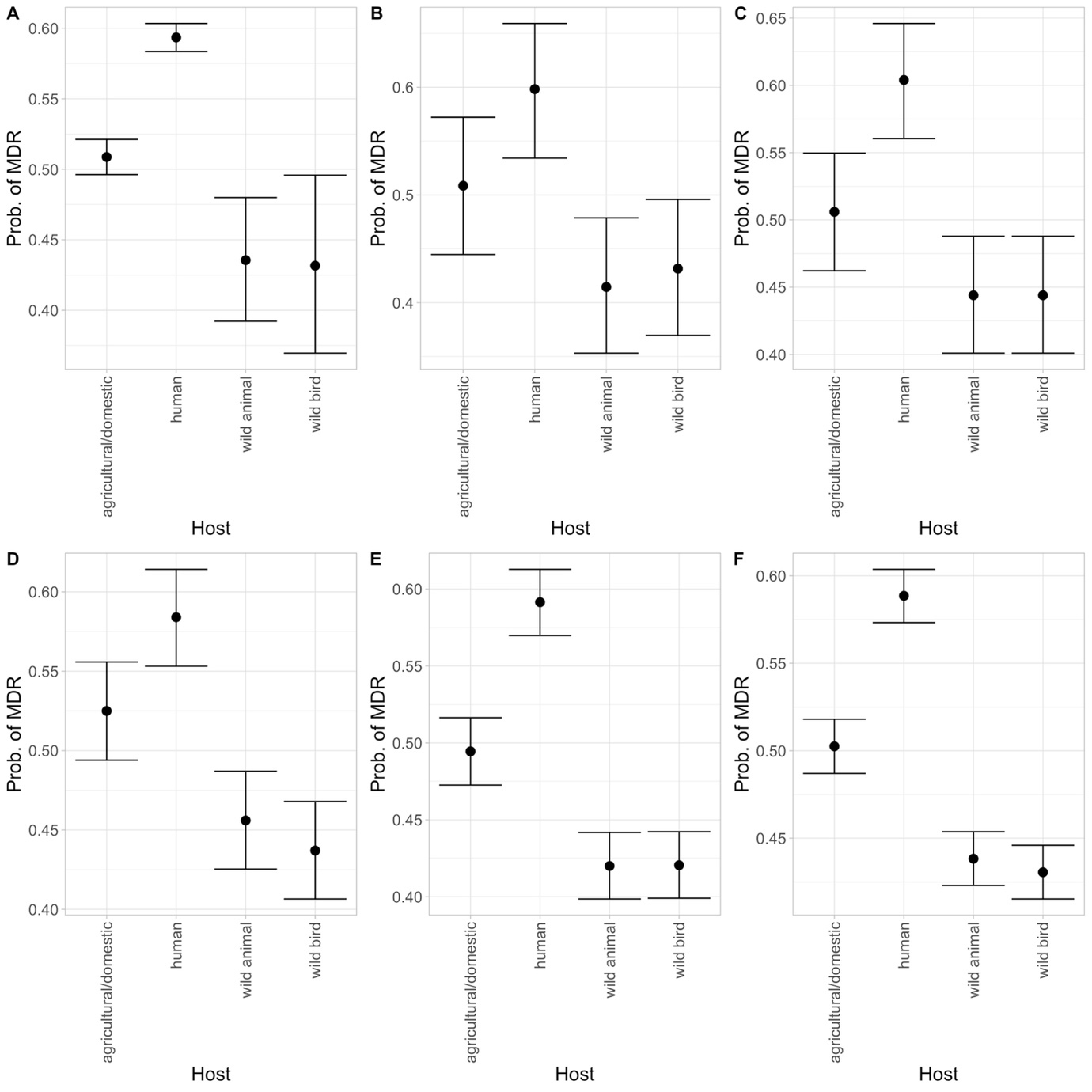

Figure S4. Estimated mean probabilities and 95% CIs of MDR according to host category from binomial GLMs using A) the full dataset B) the dataset subsampled without replacement to the minimum group size, and the dataset sampled with replacement to sample sizes of C) 500, D) 1000, E) 2000 and F) 4000.

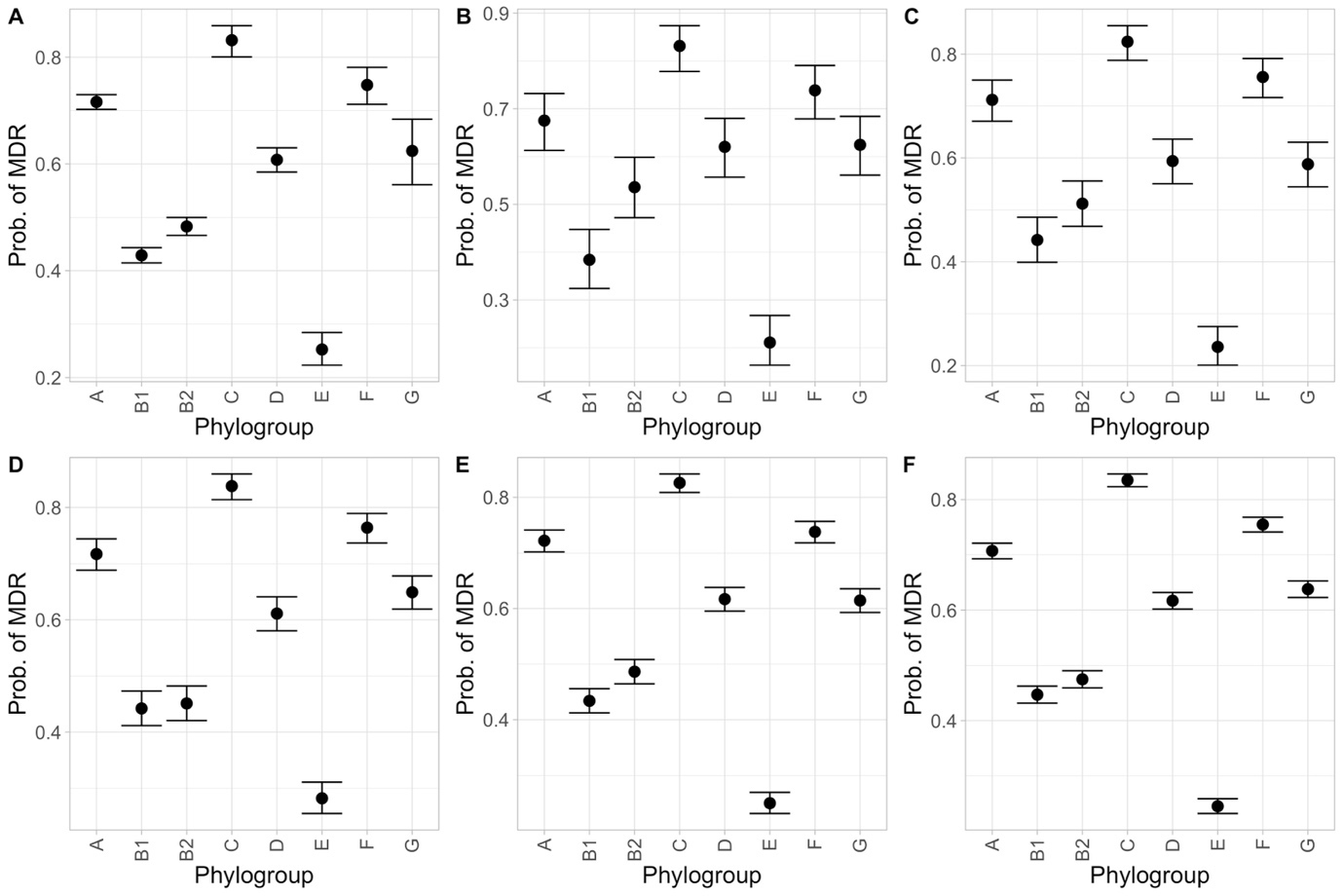

Figure S5. Estimated mean probabilities and 95% CIs of MDR according to phylogroup from binomial GLMs using A) the full dataset B) the dataset subsampled without replacement to the minimum group size, and the dataset sampled with replacement to sample sizes of C) 500, D) 1000, E) 2000 and F) 4000.

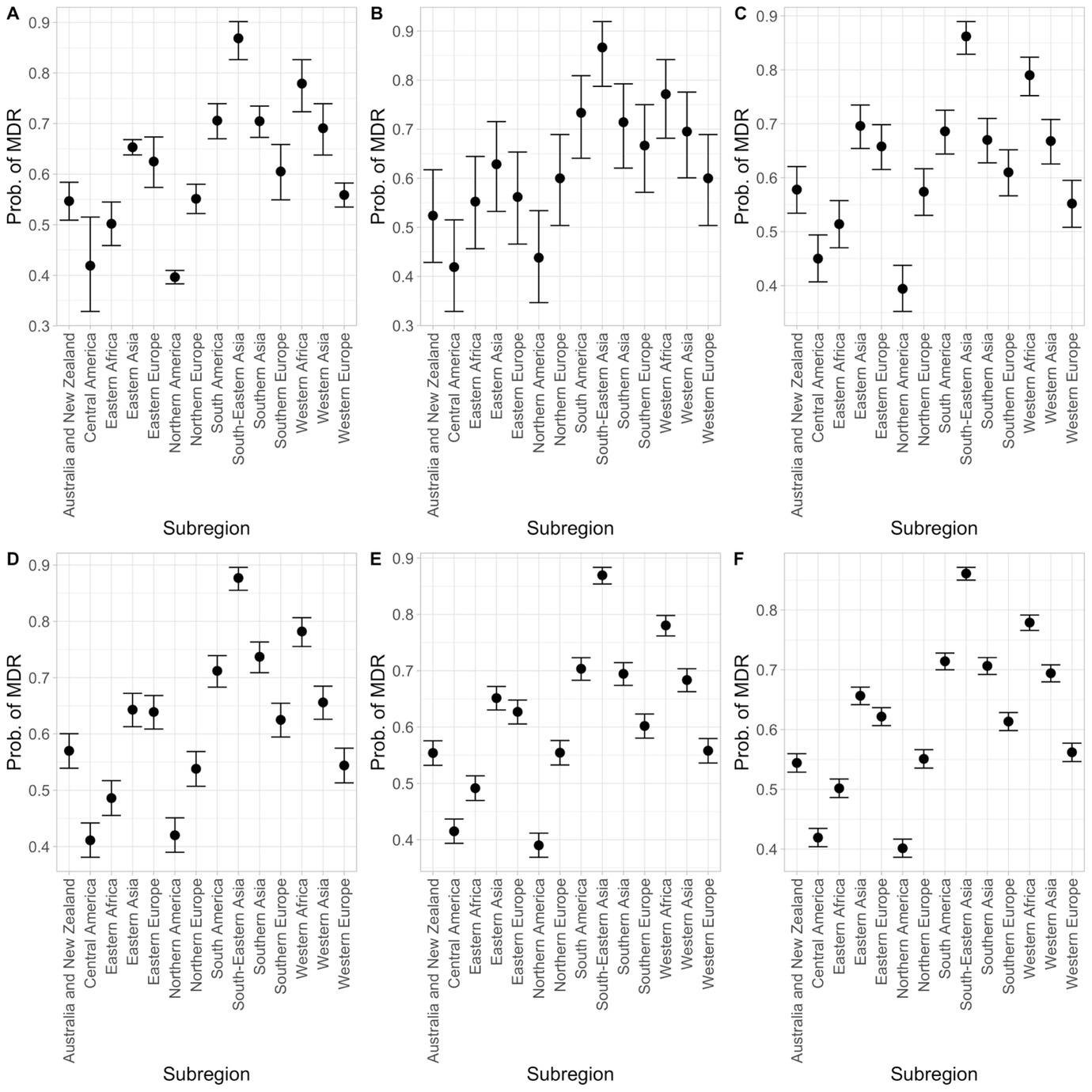

Figure S6. Estimated mean probabilities and 95% CIs of MDR according to geographic subregion from binomial GLMs using A) the full dataset B) the dataset subsampled without replacement to the minimum group size, and the dataset sampled with replacement to sample sizes of C) 500, D) 1000, E) 2000 and F) 4000.

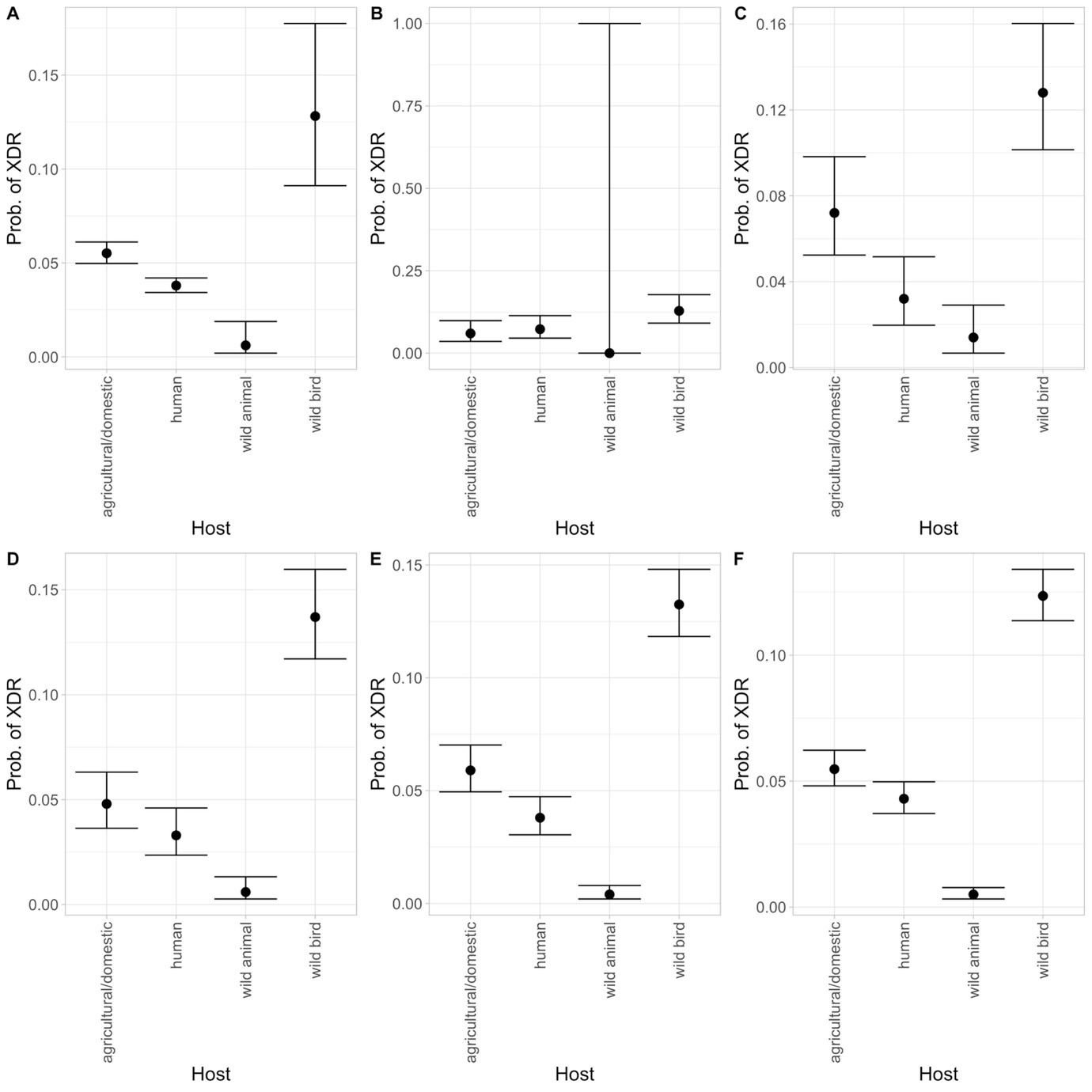

Figure S7. Estimated mean probabilities and 95% CIs of XDR according to host category from binomial GLMs using A) the full dataset B) the dataset subsampled without replacement to the minimum group size, and the dataset sampled with replacement to sample sizes of C) 500, D) 1000, E) 2000 and F) 4000. Where error bars span the full range, there was insufficient data to estimate the probability for this level of the predictor.

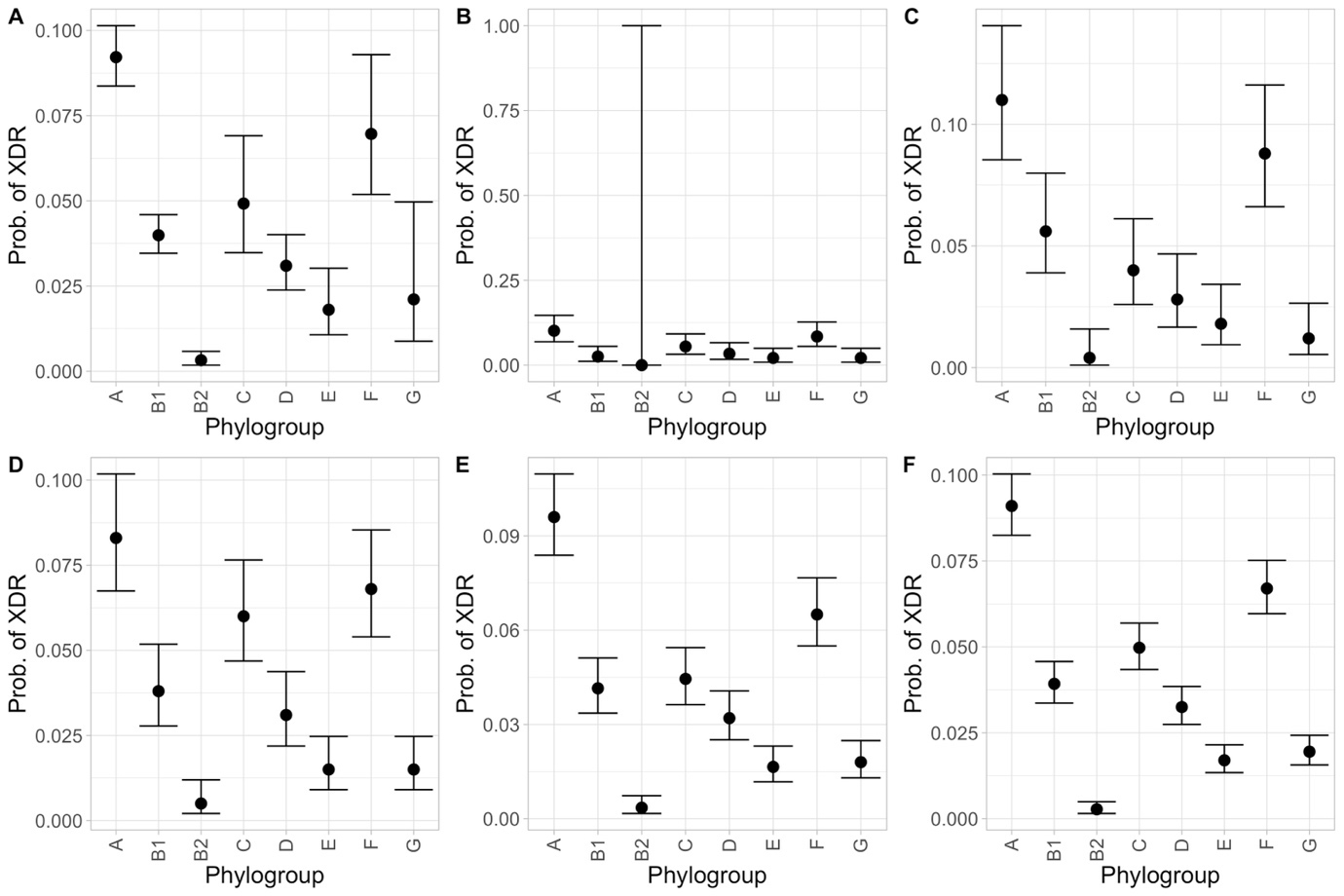

Figure S8. Estimated mean probabilities and 95% CIs of XDR according to phylogroup from binomial GLMs using A) the full dataset B) the dataset subsampled without replacement to the minimum group size, and the dataset sampled with replacement to sample sizes of C) 500, D) 1000, E) 2000 and F) 4000. Where error bars span the full range, there was insufficient data to estimate the probability for this level of the predictor.

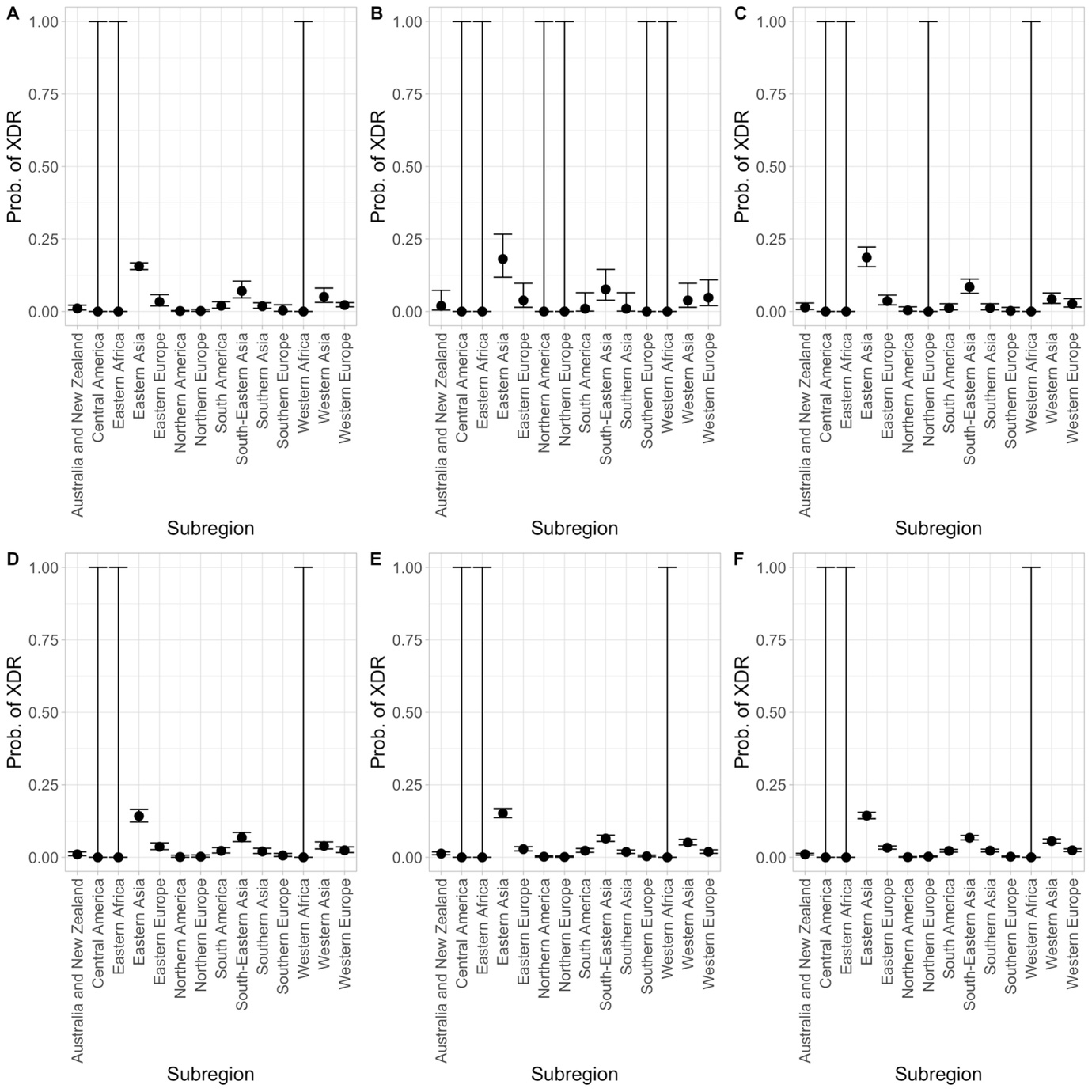

Figure S9. Estimated mean probabilities and 95% CIs of XDR according to geographic subregion from binomial GLMs using A) the full dataset B) the dataset subsampled without replacement to the minimum group size, and the dataset sampled with replacement to sample sizes of C) 500, D) 1000, E) 2000 and F) 4000. Where error bars span the full range, there was insufficient data to estimate the probability for this level of the predictor.

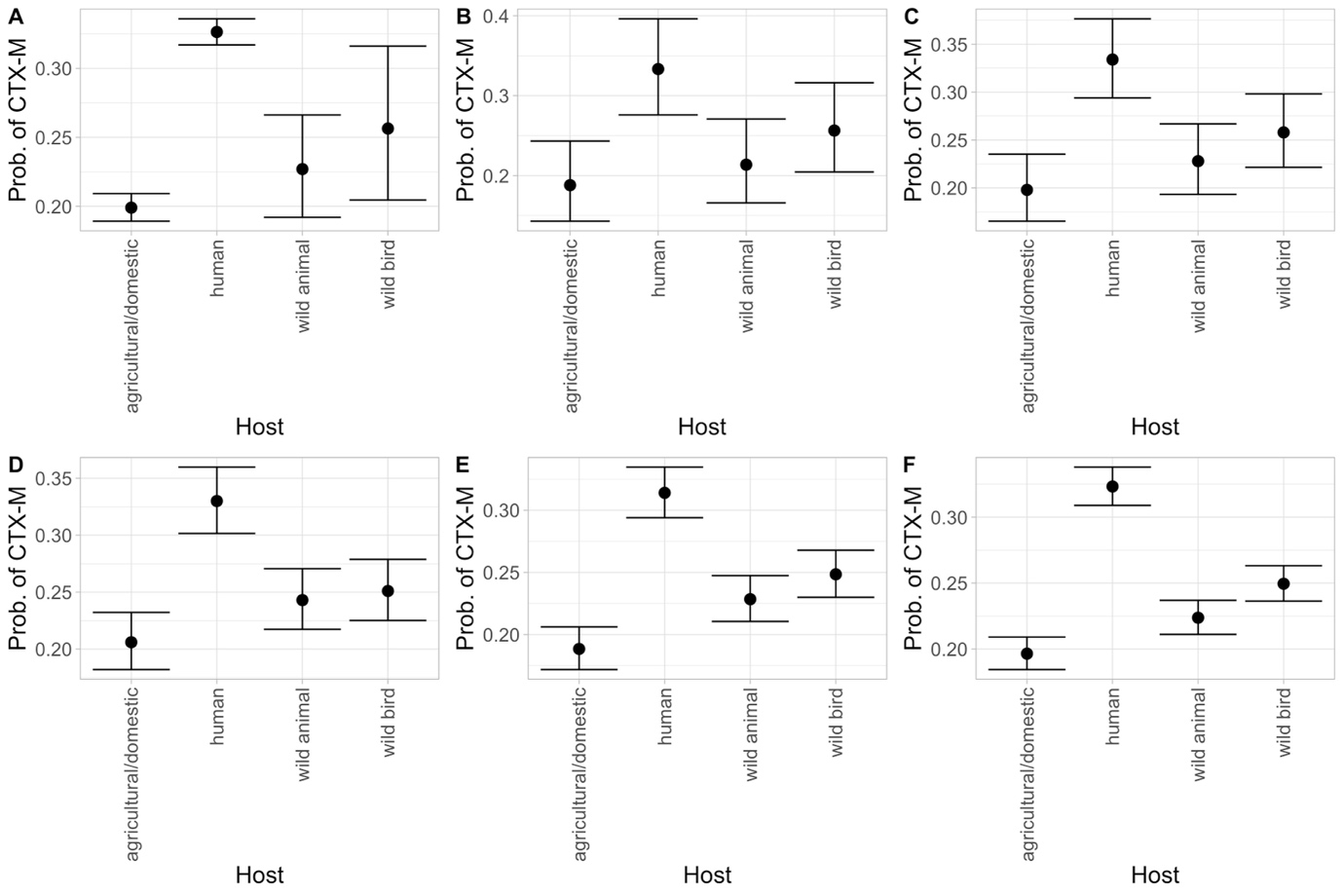

Figure S10. Estimated mean probabilities and 95% CIs of *bla*_CTX-M_ presence according to host category from binomial GLMs using A) the full dataset B) the dataset subsampled without replacement to the minimum group size, and the dataset sampled with replacement to sample sizes of C) 500, D) 1000, E) 2000 and F) 4000.

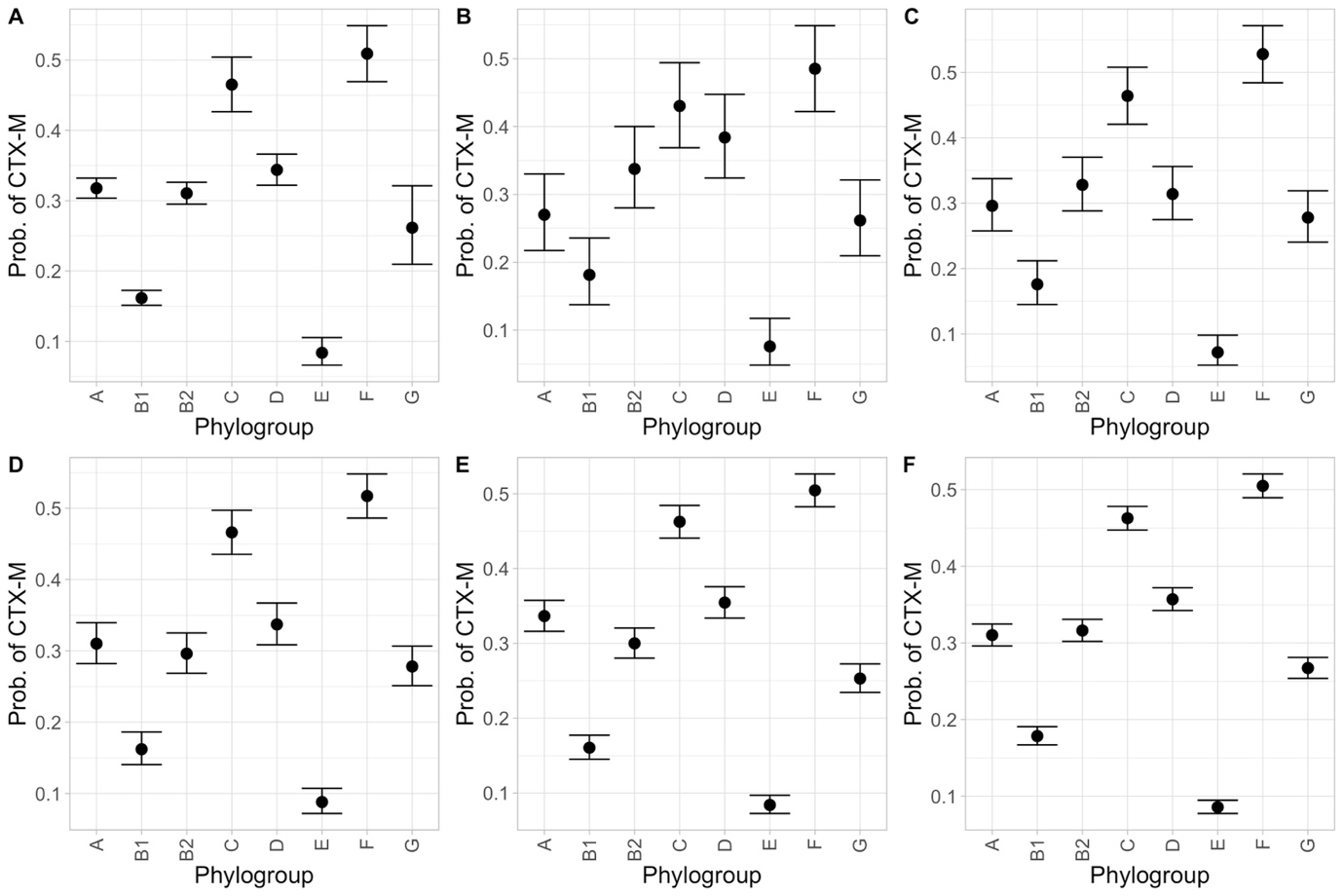

Figure S11. Estimated mean probabilities and 95% CIs of *bla*_CTX-M_ presence according to phylogroup from binomial GLMs using A) the full dataset B) the dataset subsampled without replacement to the minimum group size, and the dataset sampled with replacement to sample sizes of C) 500, D) 1000, E) 2000 and F) 4000.

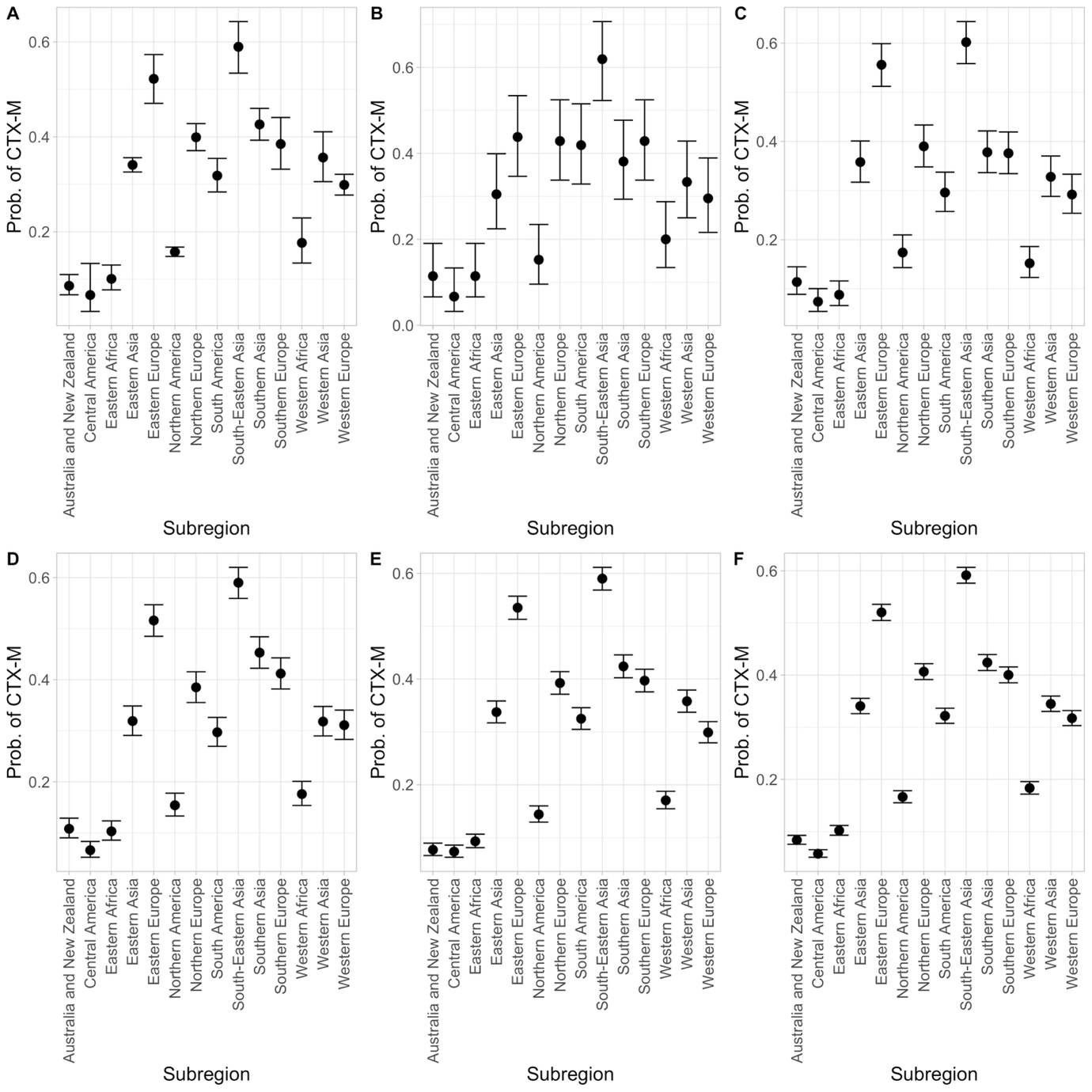

Figure S12. Estimated mean probabilities and 95% CIs of *bla*_CTX-M_ presence according to geographic subregion from binomial GLMs using A) the full dataset B) the dataset subsampled without replacement to the minimum group size, and the dataset sampled with replacement to sample sizes of C) 500, D) 1000, E) 2000 and F) 4000.

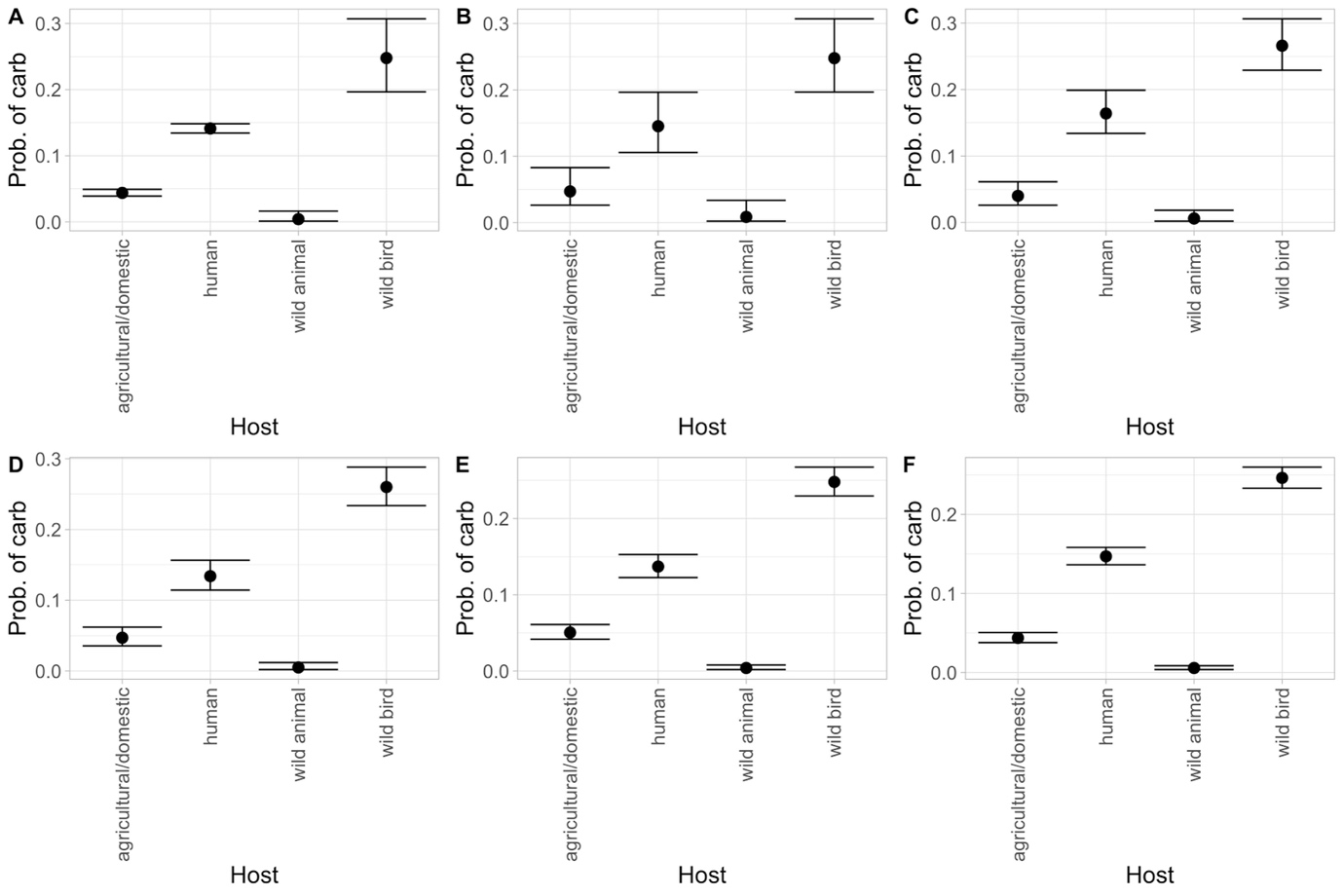

Figure S13. Estimated mean probabilities and 95% CIs of carbapenemase presence according to host category from binomial GLMs using A) the full dataset B) the dataset subsampled without replacement to the minimum group size, and the dataset sampled with replacement to sample sizes of C) 500, D) 1000, E) 2000 and F) 4000.

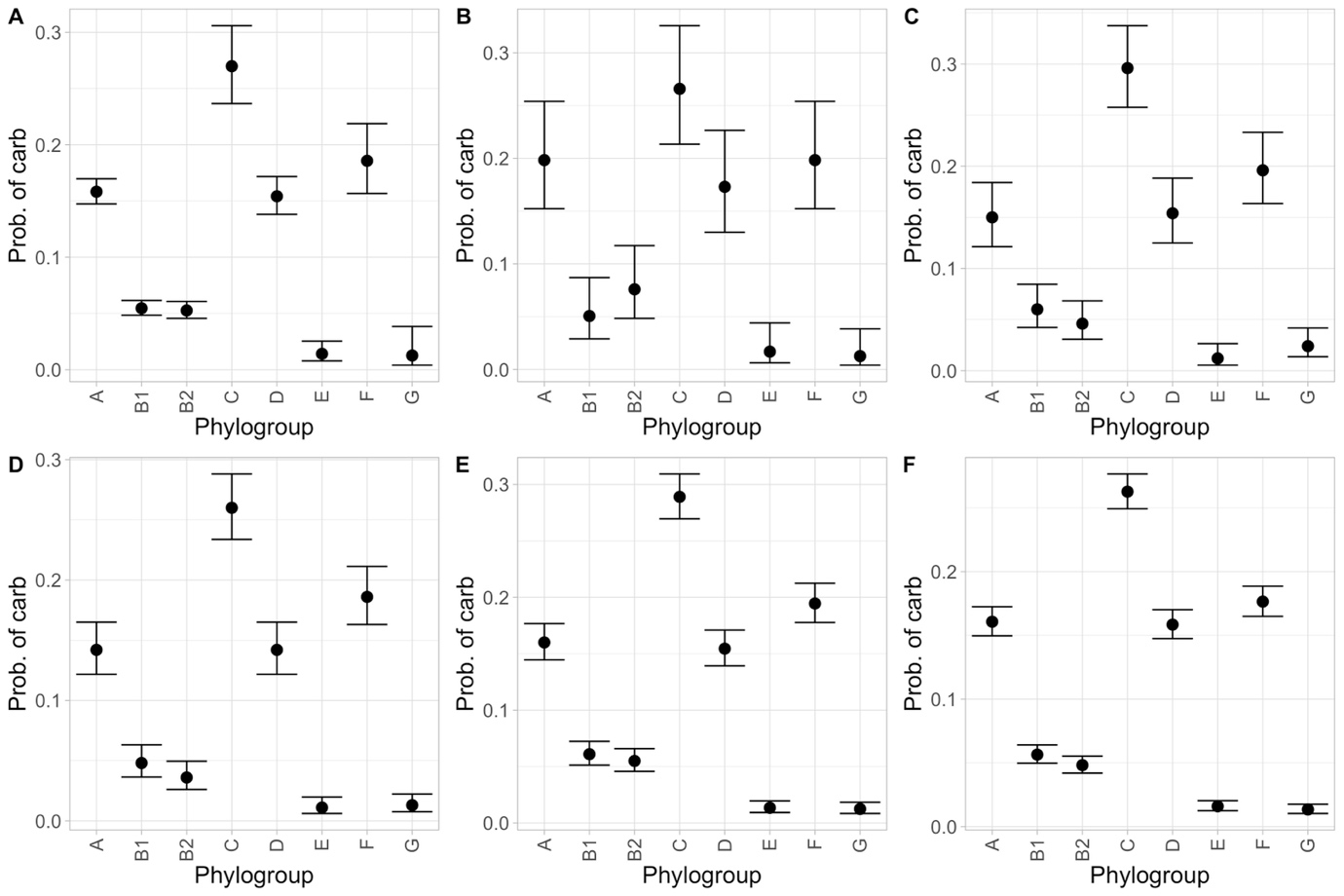

Figure S14. Estimated mean probabilities and 95% CIs of carbapenemase presence according to phylogroup from binomial GLMs using A) the full dataset B) the dataset subsampled without replacement to the minimum group size, and the dataset sampled with replacement to sample sizes of C) 500, D) 1000, E) 2000 and F) 4000.

**Supplementary Tables**

| host | count | category |
| --- | --- | --- |
| agricultural/domestic | 3126 | MDR |
| human | 5581 | MDR |
| wild animal | 213 | MDR |
| wild bird | 101 | MDR |
| agricultural/domestic | 339 | XDR |
| human | 357 | XDR |
| wild animal | 3 | XDR |
| wild bird | 30 | XDR |
| agricultural/domestic | 1223 | CTX-M |
| human | 3070 | CTX-M |
| wild animal | 111 | CTX-M |
| wild bird | 60 | CTX-M |
| agricultural/domestic | 269 | carbapenemase |
| human | 1328 | carbapenemase |
| wild animal | 2 | carbapenemase |
| wild bird | 58 | carbapenemase |

Table S1. Sample sizes of MDR, XDR, *bla*_CTX-M_ and carbapenemase genes according to host category in the full dataset.

| phylogroup | count | category |
| --- | --- | --- |
| A | 2937 | MDR |
| B1 | 1977 | MDR |
| B2 | 1641 | MDR |
| C | 524 | MDR |
| cryptic | 20 | MDR |
| D | 1080 | MDR |
| E | 196 | MDR |
| F | 451 | MDR |
| G | 148 | MDR |
| U | 39 | MDR |
| U/cryptic | 8 | MDR |
| A | 378 | XDR |
| B1 | 184 | XDR |
| B2 | 11 | XDR |
| C | 31 | XDR |
| cryptic | 3 | XDR |
| D | 55 | XDR |
| E | 14 | XDR |
| F | 42 | XDR |
| G | 5 | XDR |
| U | 5 | XDR |
| U/cryptic | 1 | XDR |
| A | 1303 | CTX-M |
| B1 | 745 | CTX-M |
| B2 | 1055 | CTX-M |
| C | 293 | CTX-M |
| cryptic | 7 | CTX-M |
| D | 611 | CTX-M |
| E | 65 | CTX-M |
| F | 307 | CTX-M |
| G | 62 | CTX-M |
| U | 15 | CTX-M |
| U/cryptic | 1 | CTX-M |
| A | 649 | carbapenemase |
| B1 | 252 | carbapenemase |
| B2 | 179 | carbapenemase |
| C | 170 | carbapenemase |
| cryptic | 1 | carbapenemase |
| D | 274 | carbapenemase |
| E | 11 | carbapenemase |
| F | 112 | carbapenemase |
| G | 3 | carbapenemase |
| U | 6 | carbapenemase |

Table S2. Sample sizes of MDR, XDR, *bla*_CTX-M_ and carbapenemase genes according to phylogroup in the full dataset.

| subregion | count | category |
| --- | --- | --- |
| Australia and New Zealand | 368 | MDR |
| Caribbean | 1 | MDR |
| Central America | 44 | MDR |
| Central Asia | 3 | MDR |
| Eastern Africa | 259 | MDR |
| Eastern Asia | 2476 | MDR |
| Eastern Europe | 225 | MDR |
| Middle Africa | 8 | MDR |
| Northern Africa | 53 | MDR |
| Northern America | 2062 | MDR |
| Northern Europe | 623 | MDR |
| South America | 468 | MDR |
| South-Eastern Asia | 271 | MDR |
| Southern Africa | 37 | MDR |
| Southern Asia | 587 | MDR |
| Southern Europe | 184 | MDR |
| Western Africa | 194 | MDR |
| Western Asia | 219 | MDR |
| Western Europe | 939 | MDR |
| Australia and New Zealand | 7 | XDR |
| Eastern Asia | 590 | XDR |
| Eastern Europe | 12 | XDR |
| Middle Africa | 1 | XDR |
| Northern Africa | 4 | XDR |
| Northern America | 8 | XDR |
| Northern Europe | 2 | XDR |
| South America | 13 | XDR |
| South-Eastern Asia | 22 | XDR |
| Southern Africa | 1 | XDR |
| Southern Asia | 15 | XDR |
| Southern Europe | 1 | XDR |
| Western Asia | 16 | XDR |
| Western Europe | 37 | XDR |
| Australia and New Zealand | 58 | CTX-M |
| Caribbean | 1 | CTX-M |
| Central America | 7 | CTX-M |
| Central Asia | 2 | CTX-M |
| Eastern Africa | 52 | CTX-M |
| Eastern Asia | 1292 | CTX-M |
| Eastern Europe | 188 | CTX-M |
| Middle Africa | 8 | CTX-M |
| Northern Africa | 35 | CTX-M |
| Northern America | 820 | CTX-M |
| Northern Europe | 451 | CTX-M |
| South America | 211 | CTX-M |
| South-Eastern Asia | 184 | CTX-M |
| Southern Africa | 24 | CTX-M |
| Southern Asia | 355 | CTX-M |
| Southern Europe | 117 | CTX-M |
| Western Africa | 44 | CTX-M |
| Western Asia | 113 | CTX-M |
| Western Europe | 502 | CTX-M |
| Australia and New Zealand | 29 | carbapenemase |
| Eastern Africa | 6 | carbapenemase |
| Eastern Asia | 771 | carbapenemase |
| Eastern Europe | 5 | carbapenemase |
| Middle Africa | 1 | carbapenemase |
| Northern Africa | 9 | carbapenemase |
| Northern America | 81 | carbapenemase |
| Northern Europe | 31 | carbapenemase |
| South America | 103 | carbapenemase |
| South-Eastern Asia | 40 | carbapenemase |
| Southern Africa | 1 | carbapenemase |
| Southern Asia | 149 | carbapenemase |
| Southern Europe | 42 | carbapenemase |
| Western Africa | 13 | carbapenemase |
| Western Asia | 68 | carbapenemase |
| Western Europe | 308 | carbapenemase |

Table S3. Sample sizes of MDR, XDR, *bla*_CTX-M_ and carbapenemase genes according to geographic subregion in the full dataset.

| cond((Int)) | disp((Int)) | cond(host) | cond(phylogroup) | cond(subregion) | df | logLik | AICc | delta | weight | model |
| --- | --- | --- | --- | --- | --- | --- | --- | --- | --- | --- |
| 0.71294294 | + | + | + | + | 24 | -9861.055 | 19770.1852 | 0 | 1 | MDR |
| 0.69841284 | + | - | + | + | 21 | -9898.9461 | 19839.9501 | 69.7648421 | 7.09E-16 | MDR |
| 0.73318464 | + | + | + | - | 11 | -10186.336 | 20394.6893 | 624.504042 | 2.46E-136 | MDR |
| 0.92302622 | + | - | + | - | 8 | -10274.06 | 20564.1288 | 793.943591 | 3.96E-173 | MDR |
| 0.12469665 | + | + | - | + | 17 | -10432.628 | 20899.2937 | 1129.10849 | 6.56E-246 | MDR |
| 0.16300412 | + | - | - | + | 14 | -10463.641 | 20955.3076 | 1185.12239 | 4.51E-258 | MDR |
| 0.02978365 | + | + | - | - | 4 | -10920.477 | 21848.9573 | 2078.77208 | 0 | MDR |
| 0.21476351 | + | - | - | - | 1 | -10995.2 | 21992.4006 | 2222.21539 | 0 | MDR |
| -4.3338199 | + | + | + | + | 24 | -2092.3453 | 4232.76582 | 0 | 1 | XDR |
| -3.7521298 | + | - | + | + | 21 | -2125.057 | 4292.17181 | 59.405998 | 1.26E-13 | XDR |
| -4.8558117 | + | + | - | + | 17 | -2201.8976 | 4437.83352 | 205.067707 | 2.95E-45 | XDR |
| -4.541752 | + | - | - | + | 14 | -2244.6861 | 4517.39846 | 284.632642 | 1.56E-62 | XDR |
| -2.2195621 | + | + | + | - | 11 | -2674.3431 | 5370.70272 | 1137.93691 | 7.95E-248 | XDR |
| -2.2827078 | + | - | + | - | 8 | -2699.1429 | 5414.29488 | 1181.52906 | 2.72E-257 | XDR |
| -2.848381 | + | + | - | - | 4 | -2879.5064 | 5767.01528 | 1534.24947 | 0 | XDR |
| -3.0634823 | + | - | - | - | 1 | -2917.7534 | 5837.50696 | 1604.74114 | 0 | XDR |
| -2.6840001 | + | + | + | + | 24 | -8360.0505 | 16768.1761 | 0 | 1 | CTX-M |
| -2.4674801 | + | - | + | + | 21 | -8405.3158 | 16852.6894 | 84.5133728 | 4.45E-19 | CTX-M |
| -2.7004708 | + | + | - | + | 17 | -8683.398 | 17400.8344 | 632.658308 | 4.17E-138 | CTX-M |
| -2.3464372 | + | - | - | + | 14 | -8776.6485 | 17581.3233 | 813.147264 | 2.67E-177 | CTX-M |
| -1.0812806 | + | + | + | - | 11 | -8862.912 | 17747.8405 | 979.664447 | 1.86E-213 | CTX-M |
| -0.7729028 | + | - | + | - | 8 | -8959.5089 | 17935.0267 | 1166.85067 | 4.18E-254 | CTX-M |
| -1.3975001 | + | + | - | - | 4 | -9226.2416 | 18460.4856 | 1692.30958 | 0 | CTX-M |
| -0.9780801 | + | - | - | - | 1 | -9380.8601 | 18763.7204 | 1995.54439 | 0 | CTX-M |
| -2.5669219 | + | + | + | NA | 11 | -4632.2914 | 9286.59931 | 0 | 1 | carbapenemase |
| -1.6709577 | + | - | + | NA | 8 | -4928.5475 | 9873.10401 | 586.504698 | 4.39E-128 | carbapenemase |
| -3.0743919 | + | + | - | NA | 4 | -5011.4233 | 10030.8491 | 744.249774 | 2.44E-162 | carbapenemase |
| -2.1700817 | + | - | - | NA | 1 | -5285.9545 | 10573.9093 | 1287.30999 | 2.91E-280 | carbapenemase |

Table S4. AIC tables for all models of MDR, XDR, *bla*_CTX-M_ and carbapenemase gene presence/absence with host, phylogroup and subregion as predictors. Subregion was not included as a fixed effect for carbapenemase models due to lack of data.

| predictor | Estimate | Std. Error | z value | Pr(>\|z\|) | model |
| --- | --- | --- | --- | --- | --- |
| (Intercept) | 0.71294294 | 0.09025457 | 7.89924446 | 2.81E-15 | MDR |
| hosthuman | 0.27665979 | 0.03902542 | 7.08922027 | 1.35E-12 | MDR |
| hostwild animal | -0.0859625 | 0.10477261 | -0.8204675 | 0.41194963 | MDR |
| hostwild bird | -0.5307505 | 0.14817734 | -3.5818601 | 0.00034116 | MDR |
| phylogroupB1 | -1.1432541 | 0.04732166 | -24.159212 | 5.97E-129 | MDR |
| phylogroupB2 | -0.9255997 | 0.05384607 | -17.189736 | 3.17E-66 | MDR |
| phylogroupC | 0.6502258 | 0.11487784 | 5.66014984 | 1.51E-08 | MDR |
| phylogroupD | -0.4346093 | 0.06255191 | -6.947978 | 3.71E-12 | MDR |
| phylogroupE | -1.7909848 | 0.09236469 | -19.390362 | 9.31E-84 | MDR |
| phylogroupF | 0.1674327 | 0.10457468 | 1.60108264 | 0.10935862 | MDR |
| phylogroupG | -0.2776835 | 0.14302629 | -1.9414856 | 0.0521994 | MDR |
| subregionCentral America | -0.7855798 | 0.22445742 | -3.4999057 | 0.00046542 | MDR |
| subregionEastern Africa | -0.3154898 | 0.12572463 | -2.5093715 | 0.01209462 | MDR |
| subregionEastern Asia | 0.3770847 | 0.09134111 | 4.12831296 | 3.65E-05 | MDR |
| subregionEastern Europe | 0.19787481 | 0.14015748 | 1.41180348 | 0.15800783 | MDR |
| subregionNorthern America | -0.4928041 | 0.08811946 | -5.592455 | 2.24E-08 | MDR |
| subregionNorthern Europe | -0.0302497 | 0.10398292 | -0.2909105 | 0.77111976 | MDR |
| subregionSouth America | 0.6364115 | 0.12226243 | 5.20529057 | 1.94E-07 | MDR |
| subregionSouth-Eastern Asia | 1.40129896 | 0.19097284 | 7.33768724 | 2.17E-13 | MDR |
| subregionSouthern Asia | 0.55662548 | 0.1160624 | 4.79591577 | 1.62E-06 | MDR |
| subregionSouthern Europe | 0.23238157 | 0.14827887 | 1.56719277 | 0.11706964 | MDR |
| subregionWestern Africa | 1.05913199 | 0.18517247 | 5.71970561 | 1.07E-08 | MDR |
| subregionWestern Asia | 0.44572587 | 0.15238517 | 2.92499502 | 0.00344462 | MDR |
| subregionWestern Europe | -0.0503682 | 0.09862096 | -0.5107253 | 0.60954341 | MDR |
| (Intercept) | -4.3338199 | 0.41752857 | -10.379697 | 3.07E-25 | XDR |
| hosthuman | -0.2903168 | 0.08640546 | -3.3599355 | 0.00077961 | XDR |
| hostwild animal | -1.7490709 | 0.59595548 | -2.934902 | 0.00333653 | XDR |
| hostwild bird | 1.68640536 | 0.26933933 | 6.26126665 | 3.82E-10 | XDR |
| phylogroupB1 | -0.9207015 | 0.09902716 | -9.2974641 | 1.44E-20 | XDR |
| phylogroupB2 | -2.749347 | 0.31148358 | -8.8266195 | 1.08E-18 | XDR |
| phylogroupC | -0.6348829 | 0.20393176 | -3.1132126 | 0.00185063 | XDR |
| phylogroupD | -0.6219379 | 0.15718154 | -3.9568129 | 7.60E-05 | XDR |
| phylogroupE | -0.8870563 | 0.2871809 | -3.0888415 | 0.00200939 | XDR |
| phylogroupF | -0.4428049 | 0.1851875 | -2.3911166 | 0.01679722 | XDR |
| phylogroupG | -1.3153603 | 0.52092235 | -2.5250602 | 0.01156785 | XDR |
| subregionCentral America | -14.720398 | 2040.43299 | -0.0072144 | 0.99424383 | XDR |
| subregionEastern Africa | -19.256017 | 8174.9254 | -0.0023555 | 0.99812059 | XDR |
| subregionEastern Asia | 3.36320099 | 0.41589369 | 8.0866843 | 6.13E-16 | XDR |
| subregionEastern Europe | 1.90673057 | 0.50511256 | 3.77486275 | 0.0001601 | XDR |
| subregionNorthern America | -1.3199004 | 0.55734264 | -2.3682029 | 0.01787473 | XDR |
| subregionNorthern Europe | -0.974838 | 0.82058609 | -1.1879777 | 0.23484219 | XDR |
| subregionSouth America | 1.17038421 | 0.49534709 | 2.36275578 | 0.01813962 | XDR |
| subregionSouth-Eastern Asia | 2.45555649 | 0.47369444 | 5.18384065 | 2.17E-07 | XDR |
| subregionSouthern Asia | 1.08792911 | 0.49038056 | 2.21854047 | 0.026518 | XDR |
| subregionSouthern Europe | -0.3853293 | 1.0844529 | -0.3553214 | 0.72234886 | XDR |
| subregionWestern Africa | -17.194362 | 4570.36579 | -0.0037621 | 0.99699825 | XDR |
| subregionWestern Asia | 1.98275342 | 0.48766881 | 4.06577864 | 4.79E-05 | XDR |
| subregionWestern Europe | 1.33669813 | 0.44559777 | 2.9997864 | 0.00270169 | XDR |
| (Intercept) | -2.6840001 | 0.14991362 | -17.903643 | 1.10E-71 | CTX-M |
| hosthuman | 0.40541925 | 0.04418079 | 9.17636857 | 4.46E-20 | CTX-M |
| hostwild animal | 0.36955701 | 0.1223102 | 3.0214734 | 0.00251548 | CTX-M |
| hostwild bird | 0.585227 | 0.17215558 | 3.39940767 | 0.00067532 | CTX-M |
| phylogroupB1 | -0.7783673 | 0.05436983 | -14.316162 | 1.73E-46 | CTX-M |
| phylogroupB2 | 0.02985339 | 0.0566652 | 0.52683813 | 0.598306 | CTX-M |
| phylogroupC | 0.59866728 | 0.09144785 | 6.54654316 | 5.89E-11 | CTX-M |
| phylogroupD | 0.2010605 | 0.06514593 | 3.08630964 | 0.00202658 | CTX-M |
| phylogroupE | -1.3682635 | 0.13696529 | -9.9898562 | 1.69E-23 | CTX-M |
| phylogroupF | 0.85174825 | 0.09440398 | 9.02237672 | 1.84E-19 | CTX-M |
| phylogroupG | -0.0779052 | 0.15976382 | -0.4876272 | 0.62581394 | CTX-M |
| subregionCentral America | -0.2433876 | 0.41997416 | -0.57953 | 0.56223158 | CTX-M |
| subregionEastern Africa | 0.39600209 | 0.2081335 | 1.90263503 | 0.05708819 | CTX-M |
| subregionEastern Asia | 1.95003101 | 0.14940685 | 13.0518178 | 6.20E-39 | CTX-M |
| subregionEastern Europe | 2.44884505 | 0.17999566 | 13.605023 | 3.74E-42 | CTX-M |
| subregionNorthern America | 0.94923805 | 0.14949655 | 6.34956489 | 2.16E-10 | CTX-M |
| subregionNorthern Europe | 2.08142871 | 0.15766694 | 13.2014273 | 8.61E-40 | CTX-M |
| subregionSouth America | 1.76068197 | 0.16818151 | 10.4689389 | 1.20E-25 | CTX-M |
| subregionSouth-Eastern Asia | 2.7370104 | 0.1882358 | 14.5403283 | 6.73E-48 | CTX-M |
| subregionSouthern Asia | 2.24902533 | 0.16290661 | 13.8056114 | 2.36E-43 | CTX-M |
| subregionSouthern Europe | 2.04124214 | 0.18905194 | 10.797256 | 3.55E-27 | CTX-M |
| subregionWestern Africa | 0.95303848 | 0.22450816 | 4.2450059 | 2.19E-05 | CTX-M |
| subregionWestern Asia | 1.94652697 | 0.18946195 | 10.2739729 | 9.23E-25 | CTX-M |
| subregionWestern Europe | 1.68442993 | 0.1552325 | 10.8510136 | 1.97E-27 | CTX-M |
| (Intercept) | -2.5669219 | 0.07008434 | -36.626185 | 1.10E-293 | carbapenemase |
| hosthuman | 1.41790189 | 0.07176026 | 19.7588738 | 6.73E-87 | carbapenemase |
| hostwild animal | -2.2867931 | 0.71218552 | -3.2109513 | 0.00132296 | carbapenemase |
| hostwild bird | 2.0248657 | 0.1725778 | 11.7330601 | 8.63E-32 | carbapenemase |
| phylogroupB1 | -1.135901 | 0.07932717 | -14.319192 | 1.66E-46 | carbapenemase |
| phylogroupB2 | -1.6136606 | 0.09028709 | -17.87255 | 1.93E-71 | carbapenemase |
| phylogroupC | 0.63004677 | 0.10408762 | 6.05304228 | 1.42E-09 | carbapenemase |
| phylogroupD | -0.2526861 | 0.08102057 | -3.1187892 | 0.00181596 | carbapenemase |
| phylogroupE | -2.529949 | 0.3079268 | -8.2160729 | 2.10E-16 | carbapenemase |
| phylogroupF | -0.0834368 | 0.11833252 | -0.7051044 | 0.48074526 | carbapenemase |
| phylogroupG | -2.5396712 | 0.58550315 | -4.3375877 | 1.44E-05 | carbapenemase |

Table S5. Model coefficients for maximal models including host, phylogroup and subregion as predictors with MDR, XDR, *bla*_CTX-M_ and carbapenemase gene presence/absence as the response variable. Subregion was not included as a fixed effect for carbapenemase models due to lack of data. Intercept in all cases is Australia and New Zealand, phylogroup A, host agricultural/domestic.

| predictor | Estimate | Std. Error | z value | Pr(>\|z\|) | model |
| --- | --- | --- | --- | --- | --- |
| (Intercept) | 0.03482798 | 0.02551731 | 1.36487646 | 0.17229188 | MDR full dataset |
| hosthuman | 0.34350391 | 0.03304372 | 10.3954381 | 2.60E-25 | MDR full dataset |
| hostwild animal | -0.2939352 | 0.09470563 | -3.1036717 | 0.00191135 | MDR full dataset |
| hostwild bird | -0.3100593 | 0.13442815 | -2.3065057 | 0.02108239 | MDR full dataset |
| (Intercept) | 0.03418601 | 0.1307632 | 0.26143451 | 0.79375745 | MDR min |
| hosthuman | 0.36415654 | 0.18676229 | 1.9498398 | 0.05119522 | MDR min |
| hostwild animal | -0.3794598 | 0.18629969 | -2.0368245 | 0.04166763 | MDR min |
| hostwild bird | -0.3094197 | 0.1857924 | -1.6654059 | 0.09583181 | MDR min |
| (Intercept) | 0.02400118 | 0.08944916 | 0.26832204 | 0.78845144 | MDR 500 |
| hosthuman | 0.3981588 | 0.12791762 | 3.11261895 | 0.00185435 | MDR 500 |
| hostwild animal | -0.2489448 | 0.12689673 | -1.9617905 | 0.04978688 | MDR 500 |
| hostwild bird | -0.248945 | 0.12689673 | -1.9617919 | 0.04978672 | MDR 500 |
| (Intercept) | 0.10008383 | 0.06332476 | 1.58048493 | 0.11399585 | MDR 1000 |
| hosthuman | 0.23912404 | 0.09014541 | 2.65264788 | 0.00798631 | MDR 1000 |
| hostwild animal | -0.2765429 | 0.08967299 | -3.083904 | 0.00204303 | MDR 1000 |
| hostwild bird | -0.3534384 | 0.08985855 | -3.9332757 | 8.38E-05 | MDR 1000 |
| (Intercept) | -0.0220009 | 0.04472407 | -0.4919262 | 0.62277152 | MDR 2000 |
| hosthuman | 0.39217049 | 0.06379295 | 6.14755177 | 7.87E-10 | MDR 2000 |
| hostwild animal | -0.3007724 | 0.06366151 | -4.7245572 | 2.31E-06 | MDR 2000 |
| hostwild bird | -0.2987199 | 0.06365623 | -4.6927051 | 2.70E-06 | MDR 2000 |
| (Intercept) | 0.01001236 | 0.03162317 | 0.31661463 | 0.75153603 | MDR 4000 |
| hosthuman | 0.34776422 | 0.0450818 | 7.71407058 | 1.22E-14 | MDR 4000 |
| hostwild animal | -0.2582905 | 0.04489449 | -5.7532788 | 8.75E-09 | MDR 4000 |
| hostwild bird | -0.2898213 | 0.04494137 | -6.4488761 | 1.13E-10 | MDR 4000 |
| (Intercept) | -2.840647 | 0.05587564 | -50.838734 | 0 | XDR full dataset |
| hosthuman | -0.3918057 | 0.07767716 | -5.0440272 | 4.56E-07 | XDR full dataset |
| hostwild animal | -2.2469486 | 0.58181846 | -3.861941 | 0.00011249 | XDR full dataset |
| hostwild bird | 0.92372439 | 0.20336514 | 4.5421964 | 5.57E-06 | XDR full dataset |
| (Intercept) | -2.7545702 | 0.27563385 | -9.9935848 | 1.63E-23 | XDR min |
| hosthuman | 0.20788621 | 0.37337091 | 0.55678202 | 0.57767637 | XDR min |
| hostwild animal | -20.499883 | 7328.79022 | -0.0027972 | 0.99776818 | XDR min |
| hostwild bird | 0.8376476 | 0.33794868 | 2.4786237 | 0.01318904 | XDR min |
| (Intercept) | -2.5563656 | 0.17301141 | -14.775706 | 2.10E-49 | XDR 500 |
| hosthuman | -0.8531307 | 0.30740699 | -2.775248 | 0.00551596 | XDR 500 |
| hostwild animal | -1.698234 | 0.41811309 | -4.061662 | 4.87E-05 | XDR 500 |
| hostwild bird | 0.63760648 | 0.21874991 | 2.91477366 | 0.00355947 | XDR 500 |
| (Intercept) | -2.9873641 | 0.14793158 | -20.194228 | 1.10E-90 | XDR 1000 |
| hosthuman | -0.3903269 | 0.23069657 | -1.6919495 | 0.09065561 | XDR 1000 |
| hostwild animal | -2.1226141 | 0.43538086 | -4.8753041 | 1.09E-06 | XDR 1000 |
| hostwild bird | 1.14693024 | 0.17418889 | 6.58440518 | 4.57E-11 | XDR 1000 |
| (Intercept) | -2.7694056 | 0.09489955 | -29.182495 | 3.23E-187 | XDR 2000 |
| hosthuman | -0.4620228 | 0.15061066 | -3.0676635 | 0.00215739 | XDR 2000 |
| hostwild animal | -2.7480488 | 0.36675346 | -7.4929048 | 6.74E-14 | XDR 2000 |
| hostwild bird | 0.8903729 | 0.11556763 | 7.70434487 | 1.32E-14 | XDR 2000 |
| (Intercept) | -2.8486717 | 0.06950315 | -40.986223 | 0 | XDR 4000 |
| hosthuman | -0.2539318 | 0.1044312 | -2.4315707 | 0.01503351 | XDR 4000 |
| hostwild animal | -2.4446315 | 0.23469524 | -10.416196 | 2.09E-25 | XDR 4000 |
| hostwild bird | 0.88897582 | 0.08449974 | 10.5204568 | 6.95E-26 | XDR 4000 |
| (Intercept) | -1.3924075 | 0.03195044 | -43.58023 | 0 | CTX-M full dataset |
| hosthuman | 0.66815326 | 0.03878713 | 17.226158 | 1.69E-66 | CTX-M full dataset |
| hostwild animal | 0.16704406 | 0.11258481 | 1.4837176 | 0.13788385 | CTX-M full dataset |
| hostwild bird | 0.32770297 | 0.15308349 | 2.14068129 | 0.03229975 | CTX-M full dataset |
| (Intercept) | -1.4628344 | 0.16730341 | -8.7436021 | 2.26E-18 | CTX-M min |
| hosthuman | 0.76968726 | 0.2173044 | 3.54197739 | 0.00039714 | CTX-M min |
| hostwild animal | 0.1599216 | 0.23113895 | 0.69188512 | 0.48900948 | CTX-M min |
| hostwild bird | 0.39812374 | 0.22450885 | 1.77330978 | 0.07617739 | CTX-M min |
| (Intercept) | -1.3988432 | 0.1122265 | -12.464464 | 1.17E-35 | CTX-M 500 |
| hosthuman | 0.70869351 | 0.14692113 | 4.82363228 | 1.41E-06 | CTX-M 500 |
| hostwild animal | 0.1792041 | 0.15478176 | 1.15778567 | 0.24695151 | CTX-M 500 |
| hostwild bird | 0.34245459 | 0.15179637 | 2.256013 | 0.02406982 | CTX-M 500 |
| (Intercept) | -1.3492049 | 0.07819083 | -17.255283 | 1.02E-66 | CTX-M 1000 |
| hosthuman | 0.64101708 | 0.10313414 | 6.21537231 | 5.12E-10 | CTX-M 1000 |
| hostwild animal | 0.21290408 | 0.10747108 | 1.98103597 | 0.04758724 | CTX-M 1000 |
| hostwild bird | 0.25591464 | 0.10692518 | 2.39339912 | 0.01669307 | CTX-M 1000 |
| (Intercept) | -1.4597707 | 0.05717194 | -25.532992 | 8.48E-144 | CTX-M 2000 |
| hosthuman | 0.67826952 | 0.0747654 | 9.07197077 | 1.17E-19 | CTX-M 2000 |
| hostwild animal | 0.24296023 | 0.07813398 | 3.10953369 | 0.00187383 | CTX-M 2000 |
| hostwild bird | 0.35313914 | 0.07711059 | 4.57964488 | 4.66E-06 | CTX-M 2000 |
| (Intercept) | -1.4083148 | 0.039792 | -35.391905 | 2.27E-274 | CTX-M 4000 |
| hosthuman | 0.66943884 | 0.05221313 | 12.8212731 | 1.25E-37 | CTX-M 4000 |
| hostwild animal | 0.16436948 | 0.05497984 | 2.98963178 | 0.00279314 | CTX-M 4000 |
| hostwild bird | 0.30703402 | 0.05402332 | 5.68336116 | 1.32E-08 | CTX-M 4000 |
| (Intercept) | -3.0839212 | 0.0623511 | -49.460578 | 0 | carbapenemase full dataset |
| hosthuman | 1.27869823 | 0.06902532 | 18.5250602 | 1.30E-76 | carbapenemase full dataset |
| hostwild animal | -2.4112076 | 0.71129943 | -3.389863 | 0.00069928 | carbapenemase full dataset |
| hostwild bird | 1.97388058 | 0.1637402 | 12.054954 | 1.83E-33 | carbapenemase full dataset |
| (Intercept) | -3.0092767 | 0.30885823 | -9.74323 | 1.97E-22 | carbapenemase min |
| hosthuman | 1.23731986 | 0.36028484 | 3.4342823 | 0.00059413 | carbapenemase min |
| hostwild animal | -1.7443131 | 0.77440525 | -2.2524551 | 0.02429352 | carbapenemase min |
| hostwild bird | 1.89923572 | 0.34397181 | 5.52148661 | 3.36E-08 | carbapenemase min |
| (Intercept) | -3.178054 | 0.22821773 | -13.925535 | 4.43E-44 | carbapenemase 500 |
| hosthuman | 1.54929193 | 0.25820689 | 6.00019585 | 1.97E-09 | carbapenemase 500 |
| hostwild animal | -1.9319233 | 0.62243757 | -3.1038025 | 0.00191051 | carbapenemase 500 |
| hostwild bird | 2.16304126 | 0.24965362 | 8.6641696 | 4.55E-18 | carbapenemase 500 |
| (Intercept) | -3.0094673 | 0.14941858 | -20.141186 | 3.22E-90 | carbapenemase 1000 |
| hosthuman | 1.14342228 | 0.17590717 | 6.50014605 | 8.02E-11 | carbapenemase 1000 |
| hostwild animal | -2.2838375 | 0.47257898 | -4.8327106 | 1.35E-06 | carbapenemase 1000 |
| hostwild bird | 1.96349879 | 0.16590183 | 11.8353053 | 2.56E-32 | carbapenemase 1000 |
| (Intercept) | -2.9339622 | 0.10211553 | -28.731792 | 1.53E-181 | carbapenemase 2000 |
| hosthuman | 1.09352843 | 0.12106441 | 9.03261702 | 1.68E-19 | carbapenemase 2000 |
| hostwild animal | -2.5834908 | 0.3686863 | -7.0072874 | 2.43E-12 | carbapenemase 2000 |
| hostwild bird | 1.82465459 | 0.11449281 | 15.9368481 | 3.52E-57 | carbapenemase 2000 |
| (Intercept) | -3.0845277 | 0.0773028 | -39.90189 | 0 | carbapenemase 4000 |
| hosthuman | 1.32620074 | 0.08927198 | 14.8557337 | 6.39E-50 | carbapenemase 4000 |
| hostwild animal | -2.0682611 | 0.22294713 | -9.2769129 | 1.74E-20 | carbapenemase 4000 |
| hostwild bird | 1.96581426 | 0.08557234 | 22.9725418 | 8.77E-117 | carbapenemase 4000 |

Table S6. Model coefficients for models with the single predictor of host category and response variables of MDR, XDR, *bla*_CTX-M_ and carbapenemase gene presence/absence, shown for the full dataset, a dataset where all groups were subsampled without replacement to the minimum group size (min), and datasets sampled with replacement to 500, 1000, 2000 and 4000 genomes per group. Intercept in all cases is agricultural/domestic.

| predictor | Estimate | Std. Error | z value | Pr(>\|z\|) | model |
| --- | --- | --- | --- | --- | --- |
| (Intercept) | 0.18777941 | 0.07743439 | 2.42501309 | 0.01530783 | MDR full dataset |
| subregionCentral America | -0.5144681 | 0.21240723 | -2.4220837 | 0.0154318 | MDR full dataset |
| subregionEastern Africa | -0.180011 | 0.11725246 | -1.535243 | 0.12472414 | MDR full dataset |
| subregionEastern Asia | 0.44579836 | 0.0846227 | 5.26807095 | 1.38E-07 | MDR full dataset |
| subregionEastern Europe | 0.32302264 | 0.13359591 | 2.41790813 | 0.01561002 | MDR full dataset |
| subregionNorthern America | -0.6083197 | 0.08245921 | -7.3772194 | 1.62E-13 | MDR full dataset |
| subregionNorthern Europe | 0.01824261 | 0.09784479 | 0.18644435 | 0.85209632 | MDR full dataset |
| subregionSouth America | 0.68770161 | 0.11515676 | 5.97187358 | 2.35E-09 | MDR full dataset |
| subregionSouth-Eastern Asia | 1.70077266 | 0.18459814 | 9.21337903 | 3.16E-20 | MDR full dataset |
| subregionSouthern Asia | 0.68191028 | 0.10846518 | 6.28690507 | 3.24E-10 | MDR full dataset |
| subregionSouthern Europe | 0.23965495 | 0.14058511 | 1.70469649 | 0.08825104 | MDR full dataset |
| subregionWestern Africa | 1.07273807 | 0.17126717 | 6.26353579 | 3.76E-10 | MDR full dataset |
| subregionWestern Asia | 0.61630065 | 0.14410496 | 4.27674818 | 1.90E-05 | MDR full dataset |
| subregionWestern Europe | 0.04903048 | 0.09170918 | 0.53462998 | 0.59290575 | MDR full dataset |
| (Intercept) | 0.09532747 | 0.19540177 | 0.48785367 | 0.62565349 | MDR min |
| subregionCentral America | -0.4220302 | 0.27803361 | -1.5179107 | 0.12903692 | MDR min |
| subregionEastern Africa | 0.11497551 | 0.2769474 | 0.41515287 | 0.67803 | MDR min |
| subregionEastern Asia | 0.43076855 | 0.28102389 | 1.53285385 | 0.12531187 | MDR min |
| subregionEastern Europe | 0.15357563 | 0.27725471 | 0.55391533 | 0.57963679 | MDR min |
| subregionNorthern America | -0.3442336 | 0.27725473 | -1.241579 | 0.21439194 | MDR min |
| subregionNorthern Europe | 0.3101386 | 0.27904194 | 1.11144081 | 0.26637866 | MDR min |
| subregionSouth America | 0.91629988 | 0.2947605 | 3.10862504 | 0.0018796 | MDR min |
| subregionSouth-Eastern Asia | 1.77647337 | 0.34727417 | 5.11547796 | 3.13E-07 | MDR min |
| subregionSouthern Asia | 0.82098495 | 0.29128844 | 2.81846045 | 0.00482546 | MDR min |
| subregionSouthern Europe | 0.59781425 | 0.28467336 | 2.10000071 | 0.03572878 | MDR min |
| subregionWestern Africa | 1.12106985 | 0.30363507 | 3.69216189 | 0.00022236 | MDR min |
| subregionWestern Asia | 0.72941327 | 0.2883241 | 2.52983803 | 0.01141152 | MDR min |
| subregionWestern Europe | 0.3101386 | 0.27904194 | 1.11144081 | 0.26637866 | MDR min |
| (Intercept) | 0.31455744 | 0.09055126 | 3.47380523 | 0.00051313 | MDR 500 |
| subregionCentral America | -0.51523 | 0.12759444 | -4.0380286 | 5.39E-05 | MDR 500 |
| subregionEastern Africa | -0.2585548 | 0.12730202 | -2.0310341 | 0.04225153 | MDR 500 |
| subregionEastern Asia | 0.51375719 | 0.13286088 | 3.86688076 | 0.00011024 | MDR 500 |
| subregionEastern Europe | 0.33983634 | 0.13071724 | 2.59978209 | 0.0093283 | MDR 500 |
| subregionNorthern America | -0.7450878 | 0.12874784 | -5.787187 | 7.16E-09 | MDR 500 |
| subregionNorthern Europe | -0.016378 | 0.1279792 | -0.1279742 | 0.89816937 | MDR 500 |
| subregionSouth America | 0.46693587 | 0.13222874 | 3.53127369 | 0.00041356 | MDR 500 |
| subregionSouth-Eastern Asia | 1.51744089 | 0.15815318 | 9.59475402 | 8.41E-22 | MDR 500 |
| subregionSouthern Asia | 0.39364088 | 0.13132118 | 2.99754299 | 0.00272165 | MDR 500 |
| subregionSouthern Europe | 0.13275347 | 0.12886591 | 1.0301675 | 0.30293138 | MDR 500 |
| subregionWestern Africa | 1.01036423 | 0.14231992 | 7.09924677 | 1.25E-12 | MDR 500 |
| subregionWestern Asia | 0.38457254 | 0.13121573 | 2.93084165 | 0.00338045 | MDR 500 |
| subregionWestern Europe | -0.1057933 | 0.12762058 | -0.8289673 | 0.40712291 | MDR 500 |
| (Intercept) | 0.28187549 | 0.06387473 | 4.41294199 | 1.02E-05 | MDR 1000 |
| subregionCentral America | -0.6417088 | 0.09061382 | -7.0817981 | 1.42E-12 | MDR 1000 |
| subregionEastern Africa | -0.3378951 | 0.08990618 | -3.7583082 | 0.00017107 | MDR 1000 |
| subregionEastern Asia | 0.30653555 | 0.09184941 | 3.33737073 | 0.00084575 | MDR 1000 |
| subregionEastern Europe | 0.28914891 | 0.09173335 | 3.15205872 | 0.00162124 | MDR 1000 |
| subregionNorthern America | -0.6046525 | 0.0904714 | -6.683355 | 2.34E-11 | MDR 1000 |
| subregionNorthern Europe | -0.1295833 | 0.09001789 | -1.439528 | 0.15000097 | MDR 1000 |
| subregionSouth America | 0.62323588 | 0.09463977 | 6.58534875 | 4.54E-11 | MDR 1000 |
| subregionSouth-Eastern Asia | 1.68244042 | 0.11554335 | 14.5611183 | 4.96E-48 | MDR 1000 |
| subregionSouthern Asia | 0.74854351 | 0.09612013 | 7.78758346 | 6.83E-15 | MDR 1000 |
| subregionSouthern Europe | 0.22894745 | 0.09135998 | 2.5059928 | 0.01221081 | MDR 1000 |
| subregionWestern Africa | 0.99547404 | 0.09972901 | 9.98178958 | 1.83E-23 | MDR 1000 |
| subregionWestern Asia | 0.36363961 | 0.09225694 | 3.94159616 | 8.09E-05 | MDR 1000 |
| subregionWestern Europe | -0.1054175 | 0.0900622 | -1.1704961 | 0.24180137 | MDR 1000 |
| (Intercept) | 0.21683534 | 0.04498446 | 4.82022814 | 1.43E-06 | MDR 2000 |
| subregionCentral America | -0.5601716 | 0.06389932 | -8.7664733 | 1.84E-18 | MDR 2000 |
| subregionEastern Africa | -0.2508365 | 0.06343642 | -3.9541402 | 7.68E-05 | MDR 2000 |
| subregionEastern Asia | 0.40880376 | 0.06500601 | 6.28870718 | 3.20E-10 | MDR 2000 |
| subregionEastern Europe | 0.30252425 | 0.0645099 | 4.68957846 | 2.74E-06 | MDR 2000 |
| subregionNorthern America | -0.6641397 | 0.06422866 | -10.340238 | 4.63E-25 | MDR 2000 |
| subregionNorthern Europe | 0.00202787 | 0.06362113 | 0.03187413 | 0.97457243 | MDR 2000 |
| subregionSouth America | 0.64719262 | 0.0664882 | 9.73394666 | 2.16E-22 | MDR 2000 |
| subregionSouth-Eastern Asia | 1.67972962 | 0.08018807 | 20.947376 | 1.98E-97 | MDR 2000 |
| subregionSouthern Asia | 0.60440915 | 0.06618313 | 9.13237472 | 6.70E-20 | MDR 2000 |
| subregionSouthern Europe | 0.1969626 | 0.06411275 | 3.07212853 | 0.00212538 | MDR 2000 |
| subregionWestern Africa | 1.05173249 | 0.07030005 | 14.9606231 | 1.33E-50 | MDR 2000 |
| subregionWestern Asia | 0.55307261 | 0.06584006 | 8.40024522 | 4.46E-17 | MDR 2000 |
| subregionWestern Europe | 0.01620695 | 0.06364651 | 0.25464005 | 0.79900112 | MDR 2000 |
| (Intercept) | 0.17750177 | 0.0317474 | 5.59106463 | 2.26E-08 | MDR 4000 |
| subregionCentral America | -0.5033682 | 0.04510743 | -11.159318 | 6.45E-29 | MDR 4000 |
| subregionEastern Africa | -0.1704942 | 0.04480971 | -3.8048507 | 0.00014189 | MDR 4000 |
| subregionEastern Asia | 0.47023687 | 0.04600552 | 10.2213138 | 1.59E-24 | MDR 4000 |
| subregionEastern Europe | 0.3194722 | 0.04550741 | 7.02022322 | 2.22E-12 | MDR 4000 |
| subregionNorthern America | -0.5767224 | 0.04525786 | -12.743033 | 3.41E-37 | MDR 4000 |
| subregionNorthern Europe | 0.0272078 | 0.04492673 | 0.60560394 | 0.54477779 | MDR 4000 |
| subregionSouth America | 0.73861412 | 0.04725257 | 15.6311932 | 4.46E-55 | MDR 4000 |
| subregionSouth-Eastern Asia | 1.6461341 | 0.05564927 | 29.5805135 | 2.66E-192 | MDR 4000 |
| subregionSouthern Asia | 0.70093092 | 0.04704824 | 14.8981325 | 3.39E-50 | MDR 4000 |
| subregionSouthern Europe | 0.28452762 | 0.04541169 | 6.26551504 | 3.72E-10 | MDR 4000 |
| subregionWestern Africa | 1.0823376 | 0.04959875 | 21.8218715 | 1.44E-105 | MDR 4000 |
| subregionWestern Asia | 0.64255599 | 0.04675101 | 13.7442164 | 5.52E-43 | MDR 4000 |
| subregionWestern Europe | 0.07177678 | 0.04498348 | 1.59562536 | 0.11057246 | MDR 4000 |
| (Intercept) | -4.5553866 | 0.37994687 | -11.989536 | 4.03E-33 | XDR full dataset |
| subregionCentral America | -15.502787 | 2213.0052 | -0.0070053 | 0.99441062 | XDR full dataset |
| subregionEastern Africa | -19.406188 | 7028.53753 | -0.0027611 | 0.997797 | XDR full dataset |
| subregionEastern Asia | 2.86460306 | 0.38257946 | 7.48760287 | 7.01E-14 | XDR full dataset |
| subregionEastern Europe | 1.18809119 | 0.48017339 | 2.47429621 | 0.0133499 | XDR full dataset |
| subregionNorthern America | -1.9204313 | 0.51918412 | -3.6989408 | 0.0002165 | XDR full dataset |
| subregionNorthern Europe | -1.7796683 | 0.80327232 | -2.215523 | 0.02672419 | XDR full dataset |
| subregionSouth America | 0.64336381 | 0.47203934 | 1.36294532 | 0.17289974 | XDR full dataset |
| subregionSouth-Eastern Asia | 1.97654867 | 0.43961622 | 4.49607764 | 6.92E-06 | XDR full dataset |
| subregionSouthern Asia | 0.55657476 | 0.46070464 | 1.20809455 | 0.2270109 | XDR full dataset |
| subregionSouthern Europe | -1.1583429 | 1.07128735 | -1.0812626 | 0.27958032 | XDR full dataset |
| subregionWestern Africa | -17.516114 | 3932.47251 | -0.0044542 | 0.99644606 | XDR full dataset |
| subregionWestern Asia | 1.62086638 | 0.45845589 | 3.53549038 | 0.00040702 | XDR full dataset |
| subregionWestern Europe | 0.76202581 | 0.41472314 | 1.83743259 | 0.06614606 | XDR full dataset |
| (Intercept) | -3.9415894 | 0.71394144 | -5.5208861 | 3.37E-08 | XDR min |
| subregionCentral America | -20.462306 | 19437.6597 | -0.0010527 | 0.99916006 | XDR min |
| subregionEastern Africa | -20.462306 | 19437.6597 | -0.0010527 | 0.99916006 | XDR min |
| subregionEastern Asia | 2.43168027 | 0.75760933 | 3.2096757 | 0.00132885 | XDR min |
| subregionEastern Europe | 0.7127633 | 0.87727608 | 0.8124732 | 0.41652016 | XDR min |
| subregionNorthern America | -20.462306 | 19437.6597 | -0.0010527 | 0.99916006 | XDR min |
| subregionNorthern Europe | -20.462306 | 19437.6597 | -0.0010527 | 0.99916006 | XDR min |
| subregionSouth America | -0.7027989 | 1.23260903 | -0.5701718 | 0.5685612 | XDR min |
| subregionSouth-Eastern Asia | 1.4463192 | 0.80313245 | 1.80084765 | 0.0717269 | XDR min |
| subregionSouthern Asia | -0.7027989 | 1.23260903 | -0.5701718 | 0.5685612 | XDR min |
| subregionSouthern Europe | -20.462306 | 19437.6597 | -0.0010527 | 0.99916006 | XDR min |
| subregionWestern Africa | -20.462306 | 19437.6597 | -0.0010527 | 0.99916006 | XDR min |
| subregionWestern Asia | 0.71276517 | 0.87727583 | 0.81247556 | 0.41651881 | XDR min |
| subregionWestern Europe | 0.9458602 | 0.84835828 | 1.11493012 | 0.26488037 | XDR min |
| (Intercept) | -4.2546153 | 0.38064131 | -11.177492 | 5.26E-29 | XDR 500 |
| subregionCentral America | -19.734582 | 7239.40815 | -0.002726 | 0.99782497 | XDR 500 |
| subregionEastern Africa | -19.734582 | 7239.40815 | -0.002726 | 0.99782497 | XDR 500 |
| subregionEastern Asia | 2.77840176 | 0.39761475 | 6.9876727 | 2.79E-12 | XDR 500 |
| subregionEastern Europe | 0.96704503 | 0.45001992 | 2.14889385 | 0.03164282 | XDR 500 |
| subregionNorthern America | -1.2628384 | 0.80429854 | -1.5701115 | 0.11638916 | XDR 500 |
| subregionNorthern Europe | -19.734582 | 7239.40815 | -0.002726 | 0.99782497 | XDR 500 |
| subregionSouth America | -0.1561636 | 0.55998142 | -0.2788728 | 0.78034248 | XDR 500 |
| subregionSouth-Eastern Asia | 1.86541606 | 0.41337722 | 4.51262428 | 6.40E-06 | XDR 500 |
| subregionSouthern Asia | -0.1561535 | 0.55997994 | -0.2788555 | 0.78035569 | XDR 500 |
| subregionSouthern Europe | -1.9579826 | 1.07092645 | -1.8283073 | 0.06750345 | XDR 500 |
| subregionWestern Africa | -19.734582 | 7239.40815 | -0.002726 | 0.99782497 | XDR 500 |
| subregionWestern Asia | 1.12743673 | 0.44112872 | 2.5557999 | 0.0105944 | XDR 500 |
| subregionWestern Europe | 0.63130145 | 0.47314288 | 1.33427233 | 0.18211462 | XDR 500 |
| (Intercept) | -4.5951198 | 0.31782083 | -14.458208 | 2.23E-47 | XDR 1000 |
| subregionCentral America | -19.404016 | 5144.53444 | -0.0037718 | 0.99699057 | XDR 1000 |
| subregionEastern Africa | -19.404016 | 5144.53444 | -0.0037718 | 0.99699057 | XDR 1000 |
| subregionEastern Asia | 2.79634273 | 0.33048121 | 8.46142722 | 2.64E-17 | XDR 1000 |
| subregionEastern Europe | 1.30754741 | 0.36031264 | 3.62892465 | 0.0002846 | XDR 1000 |
| subregionNorthern America | -2.311635 | 1.04976708 | -2.2020456 | 0.02766209 | XDR 1000 |
| subregionNorthern Europe | -1.6174863 | 0.77589432 | -2.0846735 | 0.03709895 | XDR 1000 |
| subregionSouth America | 0.80065256 | 0.38404051 | 2.08481279 | 0.0370863 | XDR 1000 |
| subregionSouth-Eastern Asia | 1.97729465 | 0.34174394 | 5.7858953 | 7.21E-09 | XDR 1000 |
| subregionSouthern Asia | 0.70329943 | 0.38991086 | 1.80374413 | 0.07127143 | XDR 1000 |
| subregionSouthern Europe | -0.5148579 | 0.51834615 | -0.9932703 | 0.32057823 | XDR 1000 |
| subregionWestern Africa | -19.404016 | 5144.53444 | -0.0037718 | 0.99699057 | XDR 1000 |
| subregionWestern Asia | 1.39070703 | 0.35733973 | 3.89183435 | 9.95E-05 | XDR 1000 |
| subregionWestern Europe | 0.88971095 | 0.37907958 | 2.34702944 | 0.01892376 | XDR 1000 |
| (Intercept) | -4.3694479 | 0.20126184 | -21.710266 | 1.64E-104 | XDR 2000 |
| subregionCentral America | -19.120072 | 2819.48317 | -0.0067814 | 0.99458926 | XDR 2000 |
| subregionEastern Africa | -19.120072 | 2819.48317 | -0.0067814 | 0.99458926 | XDR 2000 |
| subregionEastern Asia | 2.64656359 | 0.21070342 | 12.5606105 | 3.48E-36 | XDR 2000 |
| subregionEastern Europe | 0.82229716 | 0.24264761 | 3.38885337 | 0.00070186 | XDR 2000 |
| subregionNorthern America | -1.843158 | 0.53945087 | -3.41673 | 0.00063378 | XDR 2000 |
| subregionNorthern Europe | -2.5373066 | 0.73553153 | -3.4496232 | 0.00056137 | XDR 2000 |
| subregionSouth America | 0.59796573 | 0.25147572 | 2.37782689 | 0.017415 | XDR 2000 |
| subregionSouth-Eastern Asia | 1.69503264 | 0.22089077 | 7.67362363 | 1.67E-14 | XDR 2000 |
| subregionSouthern Asia | 0.37022689 | 0.26228478 | 1.41154546 | 0.15808384 | XDR 2000 |
| subregionSouthern Europe | -1.2820367 | 0.42879481 | -2.9898607 | 0.00279105 | XDR 2000 |
| subregionWestern Africa | -19.120072 | 2819.48317 | -0.0067814 | 0.99458926 | XDR 2000 |
| subregionWestern Asia | 1.44586418 | 0.22547089 | 6.41264231 | 1.43E-10 | XDR 2000 |
| subregionWestern Europe | 0.3981367 | 0.2608501 | 1.52630457 | 0.12693403 | XDR 2000 |
| (Intercept) | -4.5951194 | 0.15891038 | -28.91642 | 7.42E-184 | XDR 4000 |
| subregionCentral America | -19.286072 | 2424.9614 | -0.0079531 | 0.99365437 | XDR 4000 |
| subregionEastern Africa | -19.286072 | 2424.9614 | -0.0079531 | 0.99365437 | XDR 4000 |
| subregionEastern Asia | 2.80859996 | 0.16518643 | 17.0026074 | 7.85E-65 | XDR 4000 |
| subregionEastern Europe | 1.20956494 | 0.18205649 | 6.64389917 | 3.05E-11 | XDR 4000 |
| subregionNorthern America | -3.0052815 | 0.72491504 | -4.1457017 | 3.39E-05 | XDR 4000 |
| subregionNorthern Europe | -1.4994527 | 0.36961346 | -4.0568129 | 4.97E-05 | XDR 4000 |
| subregionSouth America | 0.78896772 | 0.19235987 | 4.10151924 | 4.10E-05 | XDR 4000 |
| subregionSouth-Eastern Asia | 1.96140682 | 0.17103159 | 11.4680966 | 1.91E-30 | XDR 4000 |
| subregionSouthern Asia | 0.82363616 | 0.1913619 | 4.30407604 | 1.68E-05 | XDR 4000 |
| subregionSouthern Europe | -1.6174862 | 0.38794707 | -4.1693477 | 3.05E-05 | XDR 4000 |
| subregionWestern Africa | -19.286072 | 2424.9614 | -0.0079531 | 0.99365437 | XDR 4000 |
| subregionWestern Asia | 1.76084704 | 0.17326772 | 10.1625799 | 2.91E-24 | XDR 4000 |
| subregionWestern Europe | 0.87898353 | 0.18982861 | 4.6304059 | 3.65E-06 | XDR 4000 |
| (Intercept) | -2.3611817 | 0.13735877 | -17.189887 | 3.16E-66 | CTX-M full dataset |
| subregionCentral America | -0.277883 | 0.41464396 | -0.6701725 | 0.50274783 | CTX-M full dataset |
| subregionEastern Africa | 0.17254121 | 0.20063241 | 0.85998671 | 0.38979637 | CTX-M full dataset |
| subregionEastern Asia | 1.70188303 | 0.14156887 | 12.0215906 | 2.74E-33 | CTX-M full dataset |
| subregionEastern Europe | 2.45013203 | 0.17320663 | 14.1457174 | 1.98E-45 | CTX-M full dataset |
| subregionNorthern America | 0.68522406 | 0.14253123 | 4.8075363 | 1.53E-06 | CTX-M full dataset |
| subregionNorthern Europe | 1.95202976 | 0.15019148 | 12.996941 | 1.27E-38 | CTX-M full dataset |
| subregionSouth America | 1.59935542 | 0.16068343 | 9.95345599 | 2.44E-23 | CTX-M full dataset |
| subregionSouth-Eastern Asia | 2.72408801 | 0.1792058 | 15.2008923 | 3.49E-52 | CTX-M full dataset |
| subregionSouthern Asia | 2.0636883 | 0.15419593 | 13.3835461 | 7.55E-41 | CTX-M full dataset |
| subregionSouthern Europe | 1.89224602 | 0.18100287 | 10.4542323 | 1.40E-25 | CTX-M full dataset |
| subregionWestern Africa | 0.82236124 | 0.2155754 | 3.81472665 | 0.00013633 | CTX-M full dataset |
| subregionWestern Asia | 1.77044067 | 0.18060727 | 9.80270993 | 1.10E-22 | CTX-M full dataset |
| subregionWestern Europe | 1.50820794 | 0.14733757 | 10.2364108 | 1.36E-24 | CTX-M full dataset |
| (Intercept) | -2.0476931 | 0.30673447 | -6.6757842 | 2.46E-11 | CTX-M min |
| subregionCentral America | -0.5913626 | 0.49713885 | -1.1895321 | 0.23423034 | CTX-M min |
| subregionEastern Africa | 1.17E-06 | 0.43378795 | 2.69E-06 | 0.99999785 | CTX-M min |
| subregionEastern Asia | 1.22296982 | 0.37287352 | 3.27985159 | 0.00103862 | CTX-M min |
| subregionEastern Europe | 1.79879726 | 0.36438211 | 4.93656854 | 7.95E-07 | CTX-M min |
| subregionNorthern America | 0.33164519 | 0.40966084 | 0.80956039 | 0.41819288 | CTX-M min |
| subregionNorthern Europe | 1.76001153 | 0.36465727 | 4.82648138 | 1.39E-06 | CTX-M min |
| subregionSouth America | 1.72100947 | 0.364975 | 4.7154174 | 2.41E-06 | CTX-M min |
| subregionSouth-Eastern Asia | 2.53320124 | 0.36670241 | 6.90805724 | 4.91E-12 | CTX-M min |
| subregionSouthern Asia | 1.56218502 | 0.36670241 | 4.2600893 | 2.04E-05 | CTX-M min |
| subregionSouthern Europe | 1.76001153 | 0.36465727 | 4.82648138 | 1.39E-06 | CTX-M min |
| subregionWestern Africa | 0.66139955 | 0.39193088 | 1.68754128 | 0.09149932 | CTX-M min |
| subregionWestern Asia | 1.35454578 | 0.37005835 | 3.66035726 | 0.00025186 | CTX-M min |
| subregionWestern Europe | 1.17761473 | 0.37397544 | 3.14890929 | 0.00163881 | CTX-M min |
| (Intercept) | -2.0505128 | 0.14071641 | -14.571952 | 4.24E-48 | CTX-M 500 |
| subregionCentral America | -0.4763001 | 0.22133245 | -2.151967 | 0.03139995 | CTX-M 500 |
| subregionEastern Africa | -0.2878077 | 0.2114751 | -1.3609533 | 0.17352845 | CTX-M 500 |
| subregionEastern Asia | 1.46644828 | 0.16882828 | 8.68603467 | 3.75E-18 | CTX-M 500 |
| subregionEastern Europe | 2.27546229 | 0.16704113 | 13.6221675 | 2.96E-42 | CTX-M 500 |
| subregionNorthern America | 0.49299194 | 0.18362053 | 2.6848411 | 0.00725643 | CTX-M 500 |
| subregionNorthern Europe | 1.60319352 | 0.16795242 | 9.54552216 | 1.35E-21 | CTX-M 500 |
| subregionSouth America | 1.18410424 | 0.17146059 | 6.90598498 | 4.99E-12 | CTX-M 500 |
| subregionSouth-Eastern Asia | 2.46432853 | 0.16777518 | 14.6882785 | 7.66E-49 | CTX-M 500 |
| subregionSouthern Asia | 1.55246121 | 0.1682485 | 9.22719178 | 2.78E-20 | CTX-M 500 |
| subregionSouthern Europe | 1.5439575 | 0.16830143 | 9.17376322 | 4.57E-20 | CTX-M 500 |
| subregionWestern Africa | 0.3315251 | 0.18792913 | 1.7640964 | 0.07771575 | CTX-M 500 |
| subregionWestern Asia | 1.33324598 | 0.16992624 | 7.84602753 | 4.29E-15 | CTX-M 500 |
| subregionWestern Europe | 1.16483119 | 0.17168356 | 6.78475652 | 1.16E-11 | CTX-M 500 |
| (Intercept) | -2.111346 | 0.10188437 | -20.722962 | 2.15E-95 | CTX-M 1000 |
| subregionCentral America | -0.5384926 | 0.16310377 | -3.301534 | 0.00096158 | CTX-M 1000 |
| subregionEastern Africa | -0.0529758 | 0.14561575 | -0.3638055 | 0.71600324 | CTX-M 1000 |
| subregionEastern Asia | 1.35298941 | 0.12240762 | 11.0531468 | 2.12E-28 | CTX-M 1000 |
| subregionEastern Europe | 2.17535727 | 0.1199355 | 18.1377263 | 1.61E-73 | CTX-M 1000 |
| subregionNorthern America | 0.40779304 | 0.13437221 | 3.03480192 | 0.00240694 | CTX-M 1000 |
| subregionNorthern Europe | 1.64295543 | 0.12084641 | 13.595401 | 4.26E-42 | CTX-M 1000 |
| subregionSouth America | 1.24971842 | 0.12316618 | 10.1466033 | 3.43E-24 | CTX-M 1000 |
| subregionSouth-Eastern Asia | 2.47528901 | 0.12047551 | 20.5459934 | 8.36E-94 | CTX-M 1000 |
| subregionSouthern Asia | 1.9227951 | 0.12006699 | 16.0143524 | 1.01E-57 | CTX-M 1000 |
| subregionSouthern Europe | 1.75563093 | 0.1204504 | 14.5755512 | 4.02E-48 | CTX-M 1000 |
| subregionWestern Africa | 0.56765885 | 0.13143758 | 4.3188473 | 1.57E-05 | CTX-M 1000 |
| subregionWestern Asia | 1.34836766 | 0.12243919 | 11.0125498 | 3.32E-28 | CTX-M 1000 |
| subregionWestern Europe | 1.3159209 | 0.12266702 | 10.7275855 | 7.55E-27 | CTX-M 1000 |
| (Intercept) | -2.4838232 | 0.08387618 | -29.612974 | 1.02E-192 | CTX-M 2000 |
| subregionCentral America | -0.0503114 | 0.11990679 | -0.4195879 | 0.67478653 | CTX-M 2000 |
| subregionEastern Africa | 0.20627587 | 0.1138545 | 1.81174991 | 0.07002485 | CTX-M 2000 |
| subregionEastern Asia | 1.8093682 | 0.09628817 | 18.7911778 | 8.92E-79 | CTX-M 2000 |
| subregionEastern Europe | 2.6240544 | 0.09510553 | 27.5909749 | 1.43E-167 | CTX-M 2000 |
| subregionNorthern America | 0.70136909 | 0.10531638 | 6.65963935 | 2.75E-11 | CTX-M 2000 |
| subregionNorthern Europe | 2.04700703 | 0.09556226 | 21.4206644 | 8.58E-102 | CTX-M 2000 |
| subregionSouth America | 1.75293382 | 0.09651123 | 18.1630034 | 1.01E-73 | CTX-M 2000 |
| subregionSouth-Eastern Asia | 2.84778668 | 0.09540536 | 29.8493356 | 8.95E-196 | CTX-M 2000 |
| subregionSouthern Asia | 2.17745286 | 0.09530222 | 22.8478706 | 1.53E-115 | CTX-M 2000 |
| subregionSouthern Europe | 2.06583666 | 0.09551884 | 21.6275298 | 9.89E-104 | CTX-M 2000 |
| subregionWestern Africa | 0.90173561 | 0.10281312 | 8.77062799 | 1.78E-18 | CTX-M 2000 |
| subregionWestern Asia | 1.89977381 | 0.09597226 | 19.7950305 | 3.29E-87 | CTX-M 2000 |
| subregionWestern Europe | 1.63175999 | 0.0970604 | 16.8118002 | 2.00E-63 | CTX-M 2000 |
| (Intercept) | -2.3957301 | 0.05715618 | -41.915507 | 0 | CTX-M 4000 |
| subregionCentral America | -0.405642 | 0.08887528 | -4.5641708 | 5.01E-06 | CTX-M 4000 |
| subregionEastern Africa | 0.21779787 | 0.07747358 | 2.81125353 | 0.00493489 | CTX-M 4000 |
| subregionEastern Asia | 1.73465418 | 0.06618248 | 26.2101706 | 2.03E-151 | CTX-M 4000 |
| subregionEastern Europe | 2.47677645 | 0.06533354 | 37.9097235 | 0 | CTX-M 4000 |
| subregionNorthern America | 0.78328653 | 0.07120703 | 11.0001293 | 3.82E-28 | CTX-M 4000 |
| subregionNorthern Europe | 2.01726487 | 0.06559777 | 30.7520361 | 1.15E-207 | CTX-M 4000 |
| subregionSouth America | 1.64998649 | 0.06642609 | 24.8394329 | 3.36E-136 | CTX-M 4000 |
| subregionSouth-Eastern Asia | 2.76590773 | 0.06558566 | 42.1724438 | 0 | CTX-M 4000 |
| subregionSouthern Asia | 2.0893588 | 0.06550174 | 31.8977585 | 2.87E-223 | CTX-M 4000 |
| subregionSouthern Europe | 1.99130471 | 0.06563748 | 30.3379213 | 3.63E-202 | CTX-M 4000 |
| subregionWestern Africa | 0.901242 | 0.07026509 | 12.8263126 | 1.17E-37 | CTX-M 4000 |
| subregionWestern Asia | 1.75352685 | 0.06613265 | 26.5152968 | 6.46E-155 | CTX-M 4000 |
| subregionWestern Europe | 1.62928108 | 0.06649076 | 24.5038723 | 1.34E-132 | CTX-M 4000 |

Table S7. Model coefficients for models with the single predictor of geographic subregion and response variables of MDR, XDR, *bla*_CTX-M_ and carbapenemase gene presence/absence, shown for the full dataset, a dataset where all groups were subsampled without replacement to the minimum group size (min), and datasets sampled with replacement to 500, 1000, 2000 and 4000 genomes per group. Intercept in all cases is Australia and New Zealand.

| predictor | Estimate | Std. Error | z value | Pr(>\|z\|) | model |
| --- | --- | --- | --- | --- | --- |
| (Intercept) | 0.92553339 | 0.03463515 | 26.7223715 | 2.59E-157 | MDR full dataset |
| phylogroupB1 | -1.2120708 | 0.04566404 | -26.543221 | 3.08E-155 | MDR full dataset |
| phylogroupB2 | -0.9938433 | 0.04876608 | -20.379807 | 2.53E-92 | MDR full dataset |
| phylogroupC | 0.67251982 | 0.11199088 | 6.00513021 | 1.91E-09 | MDR full dataset |
| phylogroupD | -0.4876024 | 0.05966775 | -8.1719593 | 3.03E-16 | MDR full dataset |
| phylogroupE | -2.010487 | 0.08958743 | -22.441618 | 1.55E-111 | MDR full dataset |
| phylogroupF | 0.16201761 | 0.0999784 | 1.62052614 | 0.1051193 | MDR full dataset |
| phylogroupG | -0.4169985 | 0.13853564 | -3.0100447 | 0.00261209 | MDR full dataset |
| (Intercept) | 0.73136844 | 0.13869756 | 5.27311667 | 1.34E-07 | MDR min |
| phylogroupB1 | -1.2041156 | 0.19254958 | -6.2535354 | 4.01E-10 | MDR min |
| phylogroupB2 | -0.5876618 | 0.19026808 | -3.0885991 | 0.00201103 | MDR min |
| phylogroupC | 0.86295604 | 0.22206567 | 3.8860399 | 0.00010189 | MDR min |
| phylogroupD | -0.2407456 | 0.19274555 | -1.2490331 | 0.21165297 | MDR min |
| phylogroupE | -2.0504541 | 0.21115068 | -9.7108573 | 2.71E-22 | MDR min |
| phylogroupF | 0.30628292 | 0.20268283 | 1.51114389 | 0.1307518 | MDR min |
| phylogroupG | -0.2227927 | 0.19295006 | -1.1546649 | 0.24822767 | MDR min |
| (Intercept) | 0.90511686 | 0.09875946 | 9.16486221 | 4.96E-20 | MDR 500 |
| phylogroupB1 | -1.1381657 | 0.13365084 | -8.5159638 | 1.65E-17 | MDR 500 |
| phylogroupB2 | -0.8571086 | 0.1332593 | -6.4318852 | 1.26E-10 | MDR 500 |
| phylogroupC | 0.63856854 | 0.15344133 | 4.16164628 | 3.16E-05 | MDR 500 |
| phylogroupD | -0.5245915 | 0.13433742 | -3.905029 | 9.42E-05 | MDR 500 |
| phylogroupE | -2.0798522 | 0.14438076 | -14.405327 | 4.79E-47 | MDR 500 |
| phylogroupF | 0.2257556 | 0.14351185 | 1.57307984 | 0.11570033 | MDR 500 |
| phylogroupG | -0.5494148 | 0.13419821 | -4.0940546 | 4.24E-05 | MDR 500 |
| (Intercept) | 0.92962142 | 0.07020152 | 13.242184 | 5.01E-40 | MDR 1000 |
| phylogroupB1 | -1.1626744 | 0.09477771 | -12.267383 | 1.36E-34 | MDR 1000 |
| phylogroupB2 | -1.1262728 | 0.09469451 | -11.89375 | 1.28E-32 | MDR 1000 |
| phylogroupC | 0.71381335 | 0.11088045 | 6.43768437 | 1.21E-10 | MDR 1000 |
| phylogroupD | -0.4780837 | 0.09558048 | -5.0018969 | 5.68E-07 | MDR 1000 |
| phylogroupE | -1.8641832 | 0.09933332 | -18.766947 | 1.41E-78 | MDR 1000 |
| phylogroupF | 0.24512769 | 0.10234491 | 2.3951137 | 0.01661521 | MDR 1000 |
| phylogroupG | -0.3149955 | 0.09653011 | -3.2631842 | 0.00110168 | MDR 1000 |
| (Intercept) | 0.95439973 | 0.04991069 | 19.1221495 | 1.65E-81 | MDR 2000 |
| phylogroupB1 | -1.2199459 | 0.06727958 | -18.132482 | 1.77E-73 | MDR 2000 |
| phylogroupB2 | -1.0084242 | 0.06702639 | -15.045182 | 3.71E-51 | MDR 2000 |
| phylogroupC | 0.60315672 | 0.07726581 | 7.80625607 | 5.89E-15 | MDR 2000 |
| phylogroupD | -0.4775842 | 0.06787433 | -7.0363019 | 1.97E-12 | MDR 2000 |
| phylogroupE | -2.0530224 | 0.07181753 | -28.586647 | 9.85E-180 | MDR 2000 |
| phylogroupF | 0.08119011 | 0.07125286 | 1.13946464 | 0.25450941 | MDR 2000 |
| phylogroupG | -0.4881223 | 0.06783634 | -7.1955875 | 6.22E-13 | MDR 2000 |
| (Intercept) | 0.88207899 | 0.03474852 | 25.3846486 | 3.73E-142 | MDR 4000 |
| phylogroupB1 | -1.0948825 | 0.04710439 | -23.243744 | 1.65E-119 | MDR 4000 |
| phylogroupB2 | -0.9831699 | 0.04701082 | -20.913696 | 4.02E-97 | MDR 4000 |
| phylogroupC | 0.74121708 | 0.05499286 | 13.4784236 | 2.10E-41 | MDR 4000 |
| phylogroupD | -0.4052441 | 0.04759609 | -8.5142313 | 1.68E-17 | MDR 4000 |
| phylogroupE | -2.0075362 | 0.05058648 | -39.685231 | 0 | MDR 4000 |
| phylogroupF | 0.24337545 | 0.05058646 | 4.81107869 | 1.50E-06 | MDR 4000 |
| phylogroupG | -0.315379 | 0.0478531 | -6.5905666 | 4.38E-11 | MDR 4000 |
| (Intercept) | -2.2873906 | 0.05398243 | -42.372875 | 0 | XDR full dataset |
| phylogroupB1 | -0.8929252 | 0.09260033 | -9.6427857 | 5.27E-22 | XDR full dataset |
| phylogroupB2 | -3.4424118 | 0.30678694 | -11.220855 | 3.22E-29 | XDR full dataset |
| phylogroupC | -0.673883 | 0.19194162 | -3.5108745 | 0.00044664 | XDR full dataset |
| phylogroupD | -1.1565206 | 0.14723004 | -7.8551943 | 3.99E-15 | XDR full dataset |
| phylogroupE | -1.7095021 | 0.27505498 | -6.2151286 | 5.13E-10 | XDR full dataset |
| phylogroupF | -0.3046629 | 0.16883781 | -1.8044711 | 0.07115748 | XDR full dataset |
| phylogroupG | -1.5499254 | 0.45522265 | -3.4047634 | 0.00066221 | XDR full dataset |
| (Intercept) | -2.1832383 | 0.21531721 | -10.139637 | 3.68E-24 | XDR min |
| phylogroupB1 | -1.46742 | 0.46621578 | -3.1475124 | 0.00164666 | XDR min |
| phylogroupB2 | -20.310004 | 4977.04132 | -0.0040807 | 0.99674405 | XDR min |
| phylogroupC | -0.6634584 | 0.35741972 | -1.8562445 | 0.06341872 | XDR min |
| phylogroupD | -1.1710421 | 0.41919959 | -2.7935193 | 0.00521379 | XDR min |
| phylogroupE | -1.6540604 | 0.50067122 | -3.3036858 | 0.00095423 | XDR min |
| phylogroupF | -0.2009268 | 0.31775744 | -0.6323275 | 0.5271729 | XDR min |
| phylogroupG | -1.6540618 | 0.5006715 | -3.3036868 | 0.00095422 | XDR min |
| (Intercept) | -2.0907414 | 0.14293009 | -14.62772 | 1.87E-48 | XDR 500 |
| phylogroupB1 | -0.7340326 | 0.24137506 | -3.0410459 | 0.00235758 | XDR 500 |
| phylogroupB2 | -3.4267045 | 0.72279557 | -4.7409041 | 2.13E-06 | XDR 500 |
| phylogroupC | -1.0873121 | 0.26928112 | -4.0378327 | 5.39E-05 | XDR 500 |
| phylogroupD | -1.4564096 | 0.3064558 | -4.7524294 | 2.01E-06 | XDR 500 |
| phylogroupE | -1.908478 | 0.36548153 | -5.221818 | 1.77E-07 | XDR 500 |
| phylogroupF | -0.2475619 | 0.21295367 | -1.1625152 | 0.24502625 | XDR 500 |
| phylogroupG | -2.3200322 | 0.43487875 | -5.3348943 | 9.56E-08 | XDR 500 |
| (Intercept) | -2.4022669 | 0.11462419 | -20.957766 | 1.59E-97 | XDR 1000 |
| phylogroupB1 | -0.8291612 | 0.20123117 | -4.120441 | 3.78E-05 | XDR 1000 |
| phylogroupB2 | -2.8910374 | 0.46275652 | -6.2474267 | 4.17E-10 | XDR 1000 |
| phylogroupC | -0.3492686 | 0.17569633 | -1.9879106 | 0.04682158 | XDR 1000 |
| phylogroupD | -1.0400106 | 0.21547334 | -4.8266326 | 1.39E-06 | XDR 1000 |
| phylogroupE | -1.7823244 | 0.2842896 | -6.2693971 | 3.62E-10 | XDR 1000 |
| phylogroupF | -0.2155582 | 0.1700516 | -1.2676048 | 0.20493911 | XDR 1000 |
| phylogroupG | -1.7823244 | 0.2842896 | -6.2693971 | 3.62E-10 | XDR 1000 |
| (Intercept) | -2.2424728 | 0.07590383 | -29.543606 | 7.93E-192 | XDR 2000 |
| phylogroupB1 | -0.89723 | 0.13539401 | -6.6268074 | 3.43E-11 | XDR 2000 |
| phylogroupB2 | -3.4090814 | 0.38617347 | -8.8278498 | 1.07E-18 | XDR 2000 |
| phylogroupC | -0.8242475 | 0.1323645 | -6.2271042 | 4.75E-10 | XDR 2000 |
| phylogroupD | -1.167025 | 0.14799643 | -7.8854946 | 3.13E-15 | XDR 2000 |
| phylogroupE | -1.8453095 | 0.19124213 | -9.6490742 | 4.96E-22 | XDR 2000 |
| phylogroupF | -0.4236878 | 0.11827285 | -3.5822914 | 0.00034059 | XDR 2000 |
| phylogroupG | -1.7567324 | 0.18452084 | -9.5205097 | 1.72E-21 | XDR 2000 |
| (Intercept) | -2.3014856 | 0.05497527 | -41.864018 | 0 | XDR 4000 |
| phylogroupB1 | -0.8962772 | 0.09824418 | -9.1229545 | 7.31E-20 | XDR 4000 |
| phylogroupB2 | -3.591915 | 0.30689093 | -11.704207 | 1.21E-31 | XDR 4000 |
| phylogroupC | -0.6482291 | 0.09116191 | -7.1107447 | 1.15E-12 | XDR 4000 |
| phylogroupD | -1.0919897 | 0.10475201 | -10.424523 | 1.92E-25 | XDR 4000 |
| phylogroupE | -1.7559102 | 0.13409878 | -13.094155 | 3.56E-39 | XDR 4000 |
| phylogroupF | -0.332227 | 0.08379484 | -3.9647666 | 7.35E-05 | XDR 4000 |
| phylogroupG | -1.6161626 | 0.12687696 | -12.738031 | 3.63E-37 | XDR 4000 |
| (Intercept) | -0.7642405 | 0.03353893 | -22.786668 | 6.22E-115 | CTX-M full dataset |
| phylogroupB1 | -0.8820914 | 0.05220989 | -16.895101 | 4.89E-64 | CTX-M full dataset |
| phylogroupB2 | -0.033652 | 0.04999531 | -0.6731041 | 0.50088109 | CTX-M full dataset |
| phylogroupC | 0.62433442 | 0.08663248 | 7.20670127 | 5.73E-13 | CTX-M full dataset |
| phylogroupD | 0.11800547 | 0.06015939 | 1.96154691 | 0.04981526 | CTX-M full dataset |
| phylogroupE | -1.628042 | 0.13385032 | -12.163154 | 4.88E-34 | CTX-M full dataset |
| phylogroupF | 0.80072914 | 0.08809409 | 9.08947675 | 9.95E-20 | CTX-M full dataset |
| phylogroupG | -0.2734051 | 0.15155235 | -1.8040306 | 0.07122651 | CTX-M full dataset |
| (Intercept) | -0.9943792 | 0.14630468 | -6.7966331 | 1.07E-11 | CTX-M min |
| phylogroupB1 | -0.5122639 | 0.22319328 | -2.2951583 | 0.02172406 | CTX-M min |
| phylogroupB2 | 0.32018494 | 0.20068466 | 1.59546291 | 0.11060876 | CTX-M min |
| phylogroupC | 0.71408018 | 0.19651052 | 3.63380137 | 0.00027928 | CTX-M min |
| phylogroupD | 0.52162437 | 0.19809952 | 2.63314307 | 0.00845987 | CTX-M min |
| phylogroupE | -1.5042937 | 0.2855266 | -5.2684887 | 1.38E-07 | CTX-M min |
| phylogroupF | 0.93528307 | 0.19569731 | 4.7792331 | 1.76E-06 | CTX-M min |
| phylogroupG | -0.0432569 | 0.20796206 | -0.2080039 | 0.83522588 | CTX-M min |
| (Intercept) | -0.8664207 | 0.09796771 | -8.8439407 | 9.24E-19 | CTX-M 500 |
| phylogroupB1 | -0.6772667 | 0.152933 | -4.4285192 | 9.49E-06 | CTX-M 500 |
| phylogroupB2 | 0.14916277 | 0.13664357 | 1.09161939 | 0.27500043 | CTX-M 500 |
| phylogroupC | 0.72216778 | 0.13281327 | 5.43746717 | 5.40E-08 | CTX-M 500 |
| phylogroupD | 0.0849283 | 0.1374139 | 0.61804738 | 0.5365441 | CTX-M 500 |
| phylogroupE | -1.6898947 | 0.19881985 | -8.4996277 | 1.90E-17 | CTX-M 500 |
| phylogroupF | 0.97854261 | 0.13275106 | 7.37126044 | 1.69E-13 | CTX-M 500 |
| phylogroupG | -0.0879734 | 0.13986407 | -0.6289921 | 0.52935425 | CTX-M 500 |
| (Intercept) | -0.8001099 | 0.06837445 | -11.701884 | 1.25E-31 | CTX-M 1000 |
| phylogroupB1 | -0.8433148 | 0.10973257 | -7.6851826 | 1.53E-14 | CTX-M 1000 |
| phylogroupB2 | -0.0663131 | 0.09733399 | -0.6812946 | 0.4956851 | CTX-M 1000 |
| phylogroupC | 0.66390281 | 0.09323973 | 7.12038564 | 1.08E-12 | CTX-M 1000 |
| phylogroupD | 0.12341543 | 0.09565941 | 1.2901545 | 0.19699702 | CTX-M 1000 |
| phylogroupE | -1.5381657 | 0.13090038 | -11.750659 | 7.01E-32 | CTX-M 1000 |
| phylogroupF | 0.86813339 | 0.09316488 | 9.31824745 | 1.18E-20 | CTX-M 1000 |
| phylogroupG | -0.1542865 | 0.09827112 | -1.5700088 | 0.11641308 | CTX-M 1000 |
| (Intercept) | -0.6789263 | 0.04732294 | -14.346664 | 1.12E-46 | CTX-M 2000 |
| phylogroupB1 | -0.9755769 | 0.07713815 | -12.647139 | 1.16E-36 | CTX-M 2000 |
| phylogroupB2 | -0.1683668 | 0.06797359 | -2.476945 | 0.01325123 | CTX-M 2000 |
| phylogroupC | 0.52864559 | 0.06519796 | 8.10831477 | 5.13E-16 | CTX-M 2000 |
| phylogroupD | 0.07961373 | 0.06651683 | 1.19689607 | 0.23134707 | CTX-M 2000 |
| phylogroupE | -1.7103013 | 0.09347644 | -18.296604 | 8.81E-75 | CTX-M 2000 |
| phylogroupF | 0.69692966 | 0.06511239 | 10.7034884 | 9.80E-27 | CTX-M 2000 |
| phylogroupG | -0.4037475 | 0.0698934 | -5.7766189 | 7.62E-09 | CTX-M 2000 |
| (Intercept) | -0.7989689 | 0.03417982 | -23.375457 | 7.60E-121 | CTX-M 4000 |
| phylogroupB1 | -0.7275754 | 0.0536017 | -13.573736 | 5.73E-42 | CTX-M 4000 |
| phylogroupB2 | 0.02790157 | 0.04821212 | 0.57872518 | 0.56277462 | CTX-M 4000 |
| phylogroupC | 0.64968269 | 0.04662448 | 13.9343689 | 3.92E-44 | CTX-M 4000 |
| phylogroupD | 0.21056239 | 0.04751149 | 4.43182024 | 9.34E-06 | CTX-M 4000 |
| phylogroupE | -1.5677019 | 0.06600884 | -23.749877 | 1.10E-124 | CTX-M 4000 |
| phylogroupF | 0.81896089 | 0.04656565 | 17.5872308 | 3.09E-69 | CTX-M 4000 |
| phylogroupG | -0.2096572 | 0.04944591 | -4.2401323 | 2.23E-05 | CTX-M 4000 |
| (Intercept) | -1.6712768 | 0.04278457 | -39.062603 | 0 | carbapenemase full dataset |
| phylogroupB1 | -1.1790645 | 0.07764176 | -15.185958 | 4.38E-52 | carbapenemase full dataset |
| phylogroupB2 | -1.2181644 | 0.08790775 | -13.857305 | 1.15E-43 | carbapenemase full dataset |
| phylogroupC | 0.67585101 | 0.09943228 | 6.79709882 | 1.07E-11 | carbapenemase full dataset |
| phylogroupD | -0.0308128 | 0.07839317 | -0.3930552 | 0.69427871 | carbapenemase full dataset |
| phylogroupE | -2.5707109 | 0.30667152 | -8.3826201 | 5.18E-17 | carbapenemase full dataset |
| phylogroupF | 0.19333265 | 0.11311827 | 1.70911962 | 0.08742879 | carbapenemase full dataset |
| phylogroupG | -2.6854384 | 0.58261429 | -4.6092904 | 4.04E-06 | carbapenemase full dataset |
| (Intercept) | -1.3968746 | 0.16291017 | -8.5745082 | 9.95E-18 | carbapenemase min |
| phylogroupB1 | -1.5343175 | 0.3381085 | -4.5379441 | 5.68E-06 | carbapenemase min |
| phylogroupB2 | -1.1018286 | 0.2943835 | -3.742834 | 0.00018196 | carbapenemase min |
| phylogroupC | 0.38095088 | 0.21945363 | 1.735906 | 0.08258046 | carbapenemase min |
| phylogroupD | -0.1676653 | 0.2367107 | -0.7083131 | 0.47875084 | carbapenemase min |
| phylogroupE | -2.6678688 | 0.52993523 | -5.0343299 | 4.80E-07 | carbapenemase min |
| phylogroupF | -2.40E-06 | 0.23038985 | -1.04E-05 | 0.9999917 | carbapenemase min |
| phylogroupG | -2.9598297 | 0.60344428 | -4.9048931 | 9.35E-07 | carbapenemase min |
| (Intercept) | -1.7346026 | 0.12524492 | -13.849684 | 1.28E-43 | carbapenemase 500 |
| phylogroupB1 | -1.0169351 | 0.22615781 | -4.4965732 | 6.91E-06 | carbapenemase 500 |
| phylogroupB2 | -1.2974184 | 0.24750945 | -5.2418943 | 1.59E-07 | carbapenemase 500 |
| phylogroupC | 0.86818567 | 0.15900927 | 5.45996881 | 4.76E-08 | carbapenemase 500 |
| phylogroupD | 0.03103904 | 0.17617416 | 0.17618385 | 0.86014951 | carbapenemase 500 |
| phylogroupE | -2.6761779 | 0.42939254 | -6.2324742 | 4.59E-10 | carbapenemase 500 |
| phylogroupF | 0.32311652 | 0.16845755 | 1.9180887 | 0.05509976 | carbapenemase 500 |
| phylogroupG | -1.9708069 | 0.31791327 | -6.1991966 | 5.68E-10 | carbapenemase 500 |
| (Intercept) | -1.7987843 | 0.09059689 | -19.854812 | 1.00E-87 | carbapenemase 1000 |
| phylogroupB1 | -1.1885807 | 0.17346921 | -6.8518252 | 7.29E-12 | carbapenemase 1000 |
| phylogroupB2 | -1.4887868 | 0.19241331 | -7.737442 | 1.01E-14 | carbapenemase 1000 |
| phylogroupC | 0.75281729 | 0.11578125 | 6.50206557 | 7.92E-11 | carbapenemase 1000 |
| phylogroupD | 8.86E-06 | 0.12812315 | 6.91E-05 | 0.99994484 | carbapenemase 1000 |
| phylogroupE | -2.7000086 | 0.31642919 | -8.5327416 | 1.43E-17 | carbapenemase 1000 |
| phylogroupF | 0.32257308 | 0.12170715 | 2.65040357 | 0.00803957 | carbapenemase 1000 |
| phylogroupG | -2.5309587 | 0.29350592 | -8.6231948 | 6.51E-18 | carbapenemase 1000 |
| (Intercept) | -1.6582299 | 0.06099379 | -27.186864 | 9.29E-163 | carbapenemase 2000 |
| phylogroupB1 | -1.0757101 | 0.11157702 | -9.6409647 | 5.37E-22 | carbapenemase 2000 |
| phylogroupB2 | -1.1856215 | 0.11549999 | -10.265122 | 1.01E-24 | carbapenemase 2000 |
| phylogroupC | 0.7579826 | 0.07844478 | 9.66262684 | 4.35E-22 | carbapenemase 2000 |
| phylogroupD | -0.0415037 | 0.08687838 | -0.4777216 | 0.63284838 | carbapenemase 2000 |
| phylogroupE | -2.6332433 | 0.20313567 | -12.962978 | 1.98E-38 | carbapenemase 2000 |
| phylogroupF | 0.2371954 | 0.08313649 | 2.85308412 | 0.00432972 | carbapenemase 2000 |
| phylogroupG | -2.7112212 | 0.21030143 | -12.892072 | 4.99E-38 | carbapenemase 2000 |
| (Intercept) | -1.6526586 | 0.04304759 | -38.391428 | 0 | carbapenemase 4000 |
| phylogroupB1 | -1.1626848 | 0.08088753 | -14.374091 | 7.53E-47 | carbapenemase 4000 |
| phylogroupB2 | -1.3292468 | 0.08542312 | -15.560738 | 1.35E-54 | carbapenemase 4000 |
| phylogroupC | 0.6209314 | 0.05606844 | 11.0745259 | 1.67E-28 | carbapenemase 4000 |
| phylogroupD | -0.0167711 | 0.061053 | -0.2746967 | 0.78354926 | carbapenemase 4000 |
| phylogroupE | -2.4663778 | 0.13316211 | -18.521619 | 1.38E-76 | carbapenemase 4000 |
| phylogroupF | 0.11241009 | 0.05977554 | 1.88053677 | 0.06003496 | carbapenemase 4000 |
| phylogroupG | -2.6388218 | 0.14361461 | -18.374327 | 2.11E-75 | carbapenemase 4000 |

Table S8. Model coefficients for models with the single predictor of phylogroup and response variables of MDR, XDR, *bla*_CTX-M_ and carbapenemase gene presence/absence, shown for the full dataset, a dataset where all groups were subsampled without replacement to the minimum group size (min), and datasets sampled with replacement to 500, 1000, 2000 and 4000 genomes per group. Intercept in all cases is phylogroup A.
